# A Phase IIa Open Label Study to Evaluate the Safety, Tolerability and Efficacy of S-1226 Administered by Nebulization in Subjects with Cystic Fibrosis Lung Disease

**DOI:** 10.1101/2021.12.10.21266937

**Authors:** Grishma Shrestha, Racheal Githumbi, Bryce Oslanski, Nadia Lachman, Daria Venkova, Ben Montgomery, Cora Pieron, John Dennis, Candice L Bjornson, Julie Jarand, Michael Parkins, Ashten Langevin, Kate Skolnik, Lori Fairservice, Clare Smith, Francis Green, Mark Montgomery

## Abstract

**Rationale:** There are approximately 35,000 people with Cystic Fibrosis (CF) in North America. This condition is characterized by impaired airway clearance resulting in chronic infection and bronchiectasis. Current airway clearance treatments include nebulized hypertonic saline and Recombinant Human DNase, which may be limited by bronchospasm and cost, respectively. S-1226, a novel biophysical therapeutic agent combines carbon dioxide (CO_2_) enriched air (a bronchodilator) with nebulized perflubron (PFOB), (a synthetic surfactant). They act synergistically to open airways, enhance mucus clearance, and increase blood oxygenation. We report preliminary results from a Phase II clinical trial.

**Methods:** An open label, single-center, Phase IIa study of subjects (≥14 years) with mild-moderate (FEV_1_ 40-80%) CF lung disease treated with multiple ascending doses of S-1226 (week one), followed by the highest tolerated dose for 5 consecutive days (week two). Each dose of S-1226 comprised three successive treatments of 3mL of perflubron nebulized (Circulaire®II) for two-minutes with CO_2_ concentrations ranging from 4 to 12%. The oxygen concentration was maintained at ambient levels. Treatments were administered twice daily. Efficacy measurements included spirometry, lung clearance index (LCI), lung volumes, blood oxygenation (SPO_2_), sputum weight and the respiratory domain of quality of life (QOL) questionnaire (CFQ-R). Safety included adverse event (AE) and tolerance monitoring, vital signs, and assessment of end-tidal CO_2_.

**Results:** We report preliminary data on safety and efficacy for six CF subjects. All subjects tolerated all doses of S-1226. There were 5 reported AEs in 3 subjects. All were mild and resolved spontaneously. End-tidal CO_2_ immediately after treatment was comparable to baseline. SpO_2_ (baseline 91-95%) rapidly improved in all subjects with treatment. Five of six subjects had improvements in their LCI. Three subjects that were compliant in collecting mucus showed increases of 14%, 29% and 64% over baseline. Percent predicted FEV_1_ response was variable, decreasing initially with return to pre-treatment values at two weeks. Four of the six subjects reported improvements in CFQ-R scores, three of which showed a clinically important difference (> 4 points). An important observation was that S-1226 controlled irritant (but not productive) cough in all five subjects and at all concentrations of S-1226.

**Conclusions:** All doses of S-1226 were safe and well tolerated. Treatments with up to 12% extrinsic CO_2_ over short periods of time did not result in an elevation of end-tidal CO_2._ The preliminary efficacy results, including overall improved oxygen saturation, CFQ-R scores, increased expectorated mucus and LCI values provide evidence for potential beneficial effects of S-1226 for CF lung disease but require larger trials and longer-term treatments to fully assess efficacy in CF.

## BACKGROUND

There are over 34,000 people living with CF in North America (Cystic Fibrosis Canada, 2018; Cystic Fibrosis Foundation, 2019). The main cause of morbidity and mortality in CF is lung disease; with respiratory failure accounting for the cause of death in 80% of CF patients (Lyczak et al., 2002).

The pathophysiology of Cystic Fibrosis (CF) lung disease involves bronchiolitis, which over time evolves into bronchiectasis. Abnormal transcellular movement of chloride ions via mutant cystic fibrosis transmembrane regulator (CFTR) proteins leads to impaired secretion clearance, increased mucus viscosity, and chronic airway inflammation. These create ideal conditions for bacterial colonization and infection (Fahy and Dickey, 2010). The airways become blocked with mucus along with bacterial aggregates (Jackson et al., 2020) and biofilm (Hoiby, 2010) resulting in impaired mucus clearance (Fahy and Dickey, 2010). The CFTR abnormality also results in an intrinsic pro-inflammatory state driven by increased phospholipase A2 production. This causes hydrolysis of unsaturated phospholipids releasing inflammatory mediators within the airway (Strandvik, 2010). In addition, studies have shown that one of the basic abnormalities in the small airways is impairment of surfactant function which leads to small airway collapse with further obstruction (Griese *et al*., 1997; Griese *et al*., 2005, Gunasekara *et al*., 2017).

Early diagnosis combined with treatment advances have dramatically increased the quality of life and survival of CF patients however, there is currently no cure. CF patients use a combination of daily therapies to manage their symptoms. Airway clearance techniques are used to loosen and clear thick mucus. Hypertonic saline and rhDNase are the primary drugs used to improve mucociliary clearance (Ratjen, 2009, Flume and Van Devanter, 2012). Current medications to manage airway obstruction, mucus clearance, and lung infection slow respiratory decline, but are not curative. Novel CFTR modulator therapies can reduce airway secretions, reduce exacerbations, and improve lung function (Habib et al 2019), however, they may be poorly tolerated (Dagenais et al 2021) and do not apply to all genetic variants. While patients may qualify for a CFTR modulator based on their genetics, cost is often prohibitive for many. Furthermore, since CFTR modulators are relatively new, most CF patients have already sustained structural airway injury and will continue to require ongoing symptomatic treatments. Also, recent studies have shown that chronic exposure to short or long-acting beta-agonists is associated with a 60% or greater reduction in F508del-CFTR activation (both wild-type and modulator corrected) (Brewington *et al*., 2018). This is of clinical significance as beta-agonists are a common addition to CF management regimen and thus may limit response to novel CFTR therapies.,

### Rationale for treating CF lung disease with S-1226

There is a clear unmet need in managing CF lung disease. Much of the pathophysiology of CF lung disease, particularly mucus retention, could potentially be mitigated with the use of S-1226. S-1226 is a new class of bronchodilator that is an aerosol/vapor/gas mixture combining pharmacological and biophysical principles for a novel mode of action. It contains a potent bronchodilator gas (extrinsic carbon dioxide or CO_2_) and nebulized perflubron, a synthetic surfactant that improves function of native surfactant and facilitates mucus clearance.

#### Potential benefits of inhaled (extrinsic) CO_2_

Extrinsic CO_2_ has been shown to relax airway smooth muscle (Duane *et al*., 1979; Twort and Cameron, 1986; El Mays *et al*., 2011) via non-adrenergic non-cholinergic pathways (Choudhury *et al*., 2012, Fisher and Hansen, 1976) and to reduce airway resistance in asthmatics (Van den Elshout et al 1991, Fisher and Hansen, 1976) Inhaled CO_2_ also improves ventilation – perfusion matching in the lungs (Brogan *et al*., 2004, Ingram 1975, Dorrington *et al*., 2010) thus enhancing arterial oxygen saturation. Inhaled CO_2_ has also been shown to attenuate acute lung injury (Tang et al., 2019), and has anti-inflammatory (Tang et al., 2019), antiviral, anti-bacterial properties (reviewed in El-Betany *et al*, 2020,) and antitussive effects (Banyai *et al*., 1944; Banyai, 1947).

#### Potential benefits of Perfluorooctylbromide (Perflubron)

Perfluorooctylbromide (Perflubron) has surfactant and mucolytic properties. It has a high density and low surface tension compatible with that of normal airways and exhibits natural surfactant-like qualities (Lehmler *et al*., 1999, Wolfson and Shaffer, 2005; Schürch *et al*., 2006). Natural surfactants enhance sputum clearance by decreasing sputum adhesivity and cohesivity (Albers *et al*., 1996, De Sanctis *et al*., 1994). They have been used successfully to treat patients with stable chronic bronchitis. After 14 days of treatment with aerosolized natural surfactant, *in vitro* mucociliary clearance of sputum and pulmonary function improved and lasted for a week after treatment cessation (Anzueto *et al*., 1997). We have shown that perflubron enhanced mucin transport in an *in vitro* model. The method measured rate of flow of pure mucin and asthmatic mucus through glass capillary tubes of a diameter equivalent to a membranous bronchiole (Al-Saiedy et al., 2013). This study also showed an additive effect when perflubron was combined with natural surfactant. In addition to its innate surfactant properties perflubron vapour has the capacity to repair dysfunctional natural surfactant (Gerber et al 2007) Perflubon, (Liquivent® Lung Lavage Solutions OriGen Biomedical) is approved for lung lavage by Health Canada. The high density of perflubron enables penetration through the bronchioles to the alveoli, thus facilitating removal of exudates (Tawfic and Kausalya, 2011)

#### Potential Benefits of S-1226

We have shown that the combination of CO_2_ and perflubron act synergistically to rapidly open collapsed and constricted airways due to its broncho-dilatory properties (El Mays *et al*., 2014). S-1226 is also expected to improve mucus clearance (Al-Saiedy et al., 2013) and blood oxygenation (SpO_2_) by opening small airways and improving ventilation to perfusion matching in diseased lungs (Brogan *et al*., 2004; Dorrington *et al*., 2010).

The above properties indicate that S-1226 would be effective for improving mucus clearance and treating CF airway disease, as well as non-CF bronchiectasis..

Thus, we undertook a Phase IIa, proof of concept open label study, to evaluate the safety and tolerability of S-1226 composed of PFOB with ascending doses of carbon dioxide (4%, 8%, and 12% CO_2_) administered in up to three successive doses, twice daily, in subjects with CF lung disease.

## METHODS

### Participants

Eligible subjects enrolled in this study had a confirmed diagnosis of CF by sweat chloride concentration over 60 mosm/L and/or genotype analysis. Additional inclusion criteria and exclusion criteria are outlined in **Table 1** below.

**Table 1:**
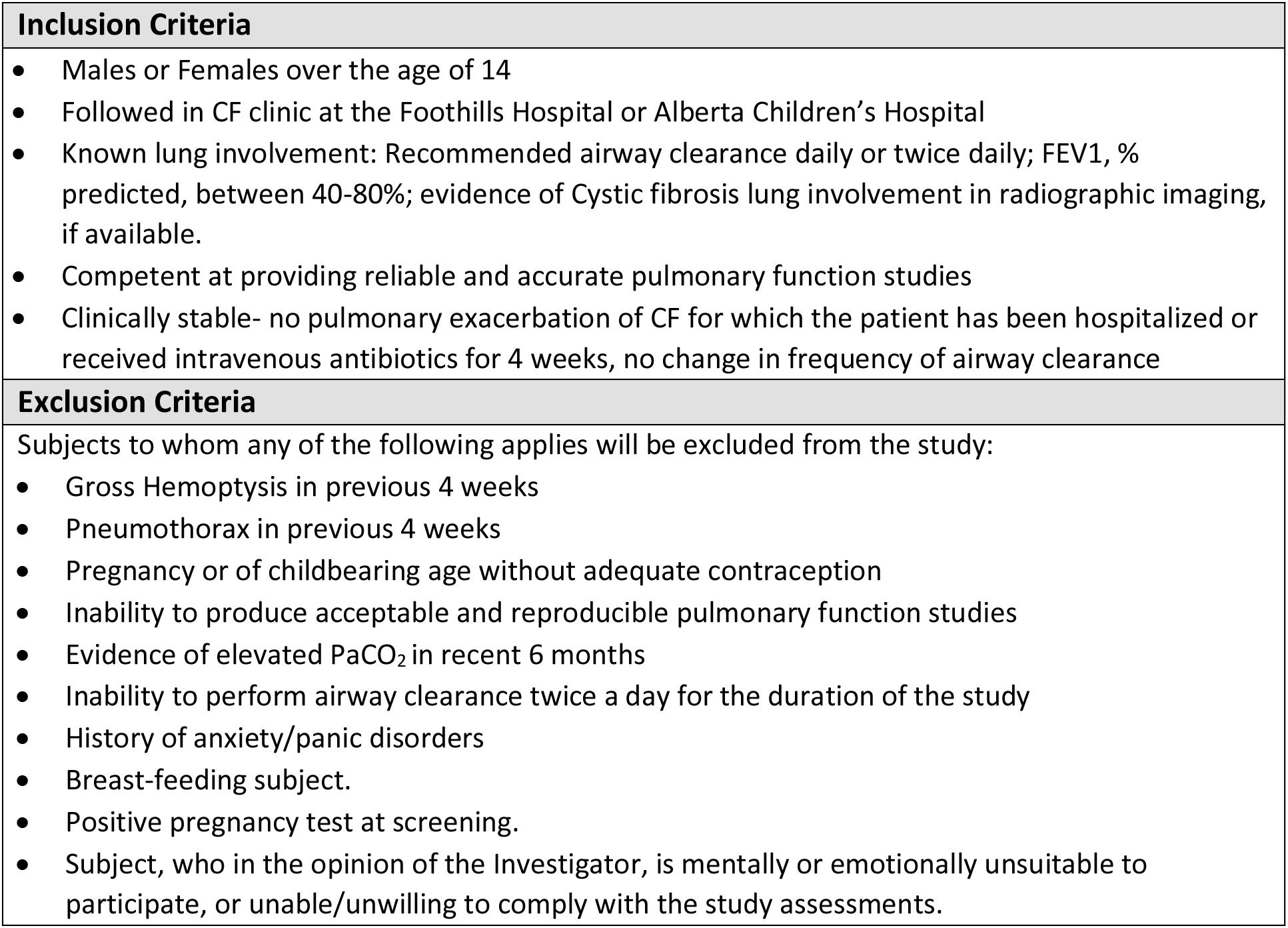
Study inclusion and Exclusion Criteria.

This study was conducted at the Alberta Lung Function Clinic, Calgary AB, from February 2019 and is currently ongoing. Procedures were conducted in accordance with the declaration of Helsinki (2014). The Conjoint Health Research Ethics Board at the University of Calgary approved the study protocol, amendment, and consent forms, (REB18-2069). Written informed consent was obtained for all subjects.

### Trial design (NCT03903913)

This is a single centre, open label, Phase IIa, multiple-ascending dose study involving subjects with mild to moderate CF lung disease. To date six subjects have been enrolled. The study consists of a screening period, a run-in, two dosing and evaluation periods (with a minimum two-day break in between) and a follow-up period. For an overview of the study schedule, refer to **Supplemental Figure A1**.

The dosing and evaluation period is divided into two consecutive components:

1. ***Dose escalation study:*** This segment of the treatment period is designed to assess the safety and tolerability of escalating doses of S-1226 (4%, 8% and 12%) in those with CF lung disease.
2. ***Daily dosing study***: This segment of the treatment period is designed to assess the short term (5 day) safety and tolerability of twice daily administration of a fixed dose of S-1226 in subjects with mild-moderate CF lung disease.

The study protocol was also amended to include a 4-week (28-day) home study extension as of October 6^th^, 2020. Regular use of adjuncts of airway clearance are generally used at home and coordinated with other CF post treatment airway clearance strategies (Mogayzel et al., 2013). These may require 1-3 treatments a day. Consequently, home use is essential for ongoing care in CF populations. The home use component of this study is an important step to demonstrate safety and tolerance for home use and involves close monitoring of blood oxygen saturation.

### Outcome measures

The primary outcome was to evaluate the safety and tolerability of S-1226 composed of PFOB with ascending doses of carbon dioxide (4%, 8%, and 12% CO_2_) administered up to three successive doses of two minutes of tidal breathing, twice daily in subjects with CF lung disease.

The secondary outcome was to evaluate the efficacy of S-1226 as measured by improvement in percent predicted Forced Expiratory Volume in 1 Second (FEV_1_) from baseline and improvement in the respiratory domain of the Cystic Fibrosis Questionnaire -Revised (CFQ-R) from baseline.

#### Other Exploratory efficacy and/or safety end points included

1. Improvement in standard lung function test parameters like Forced Expiratory Flow at 25-75% (FEF_25-75%_), Forced Vital Capacity (FVC), Inspiratory Capacity (IC), Thoracic Gas Volume (TGV) and Residual Volume (RV) from baseline
2. Improvement in Lung Clearance Index (LCI) from baseline
3. Change in expectorated sputum weight
4. Change in oxygen saturation via pulse oximetry (SpO_2_)
5. Change in end-tidal CO_2_ from baseline

### Data analyses

#### Sample Size

This exploratory study was designed to evaluate the safety and tolerability of S-1226 in up to 12 subjects with CF lung disease. A formal sample size calculation was not considered appropriate at this time as this is a short-term, small-scale study with the primary aim to gather preliminary safety and tolerability data on S-1226. S-1226 has been previously tested for safety at the doses proposed in this study in 36 healthy individuals and 12 asthmatics, with no clinically significant adverse events found (Green et al, 2016, Swystun et al, 2018). Assuming responses to S-1226 in CF subjects are not different from previous subjects, we expected to see similar safety and tolerability data. Response data from this trial will be used to inform statistical power calculations for a larger trial.

#### Safety Evaluations

Demographic parameters are summarised descriptively. Treatment-emergent adverse events are summarised descriptively by treatment for all subjects who were dosed (safety population). No inferential statistical analysis of safety data was conducted. Adverse events (AEs) were assessed by monitoring the incidence and severity of recorded AEs and summarized by group within system organ class (SOC), the Medical Dictionary for Regulatory Activities (MedDRA) and preferred terms (PTs).

Oxygen Saturation (SpO_2_) and End Tidal Carbon Dioxide (ETCO_2_) were obtained for all subjects during the baseline week and continuously monitored throughout all treatments using a Capnostream 35 Portable Respiratory Monitor. Values were obtained at each second to generate a times-series. The raw data was de-noised using exponential smoothing at an appropriate dampening factor (1 – α). This was done to remove high frequency noise within the datasets and improve pattern recognition during analysis. **Figure 1** illustrates a characteristic treatment response.

**Figure 1:**
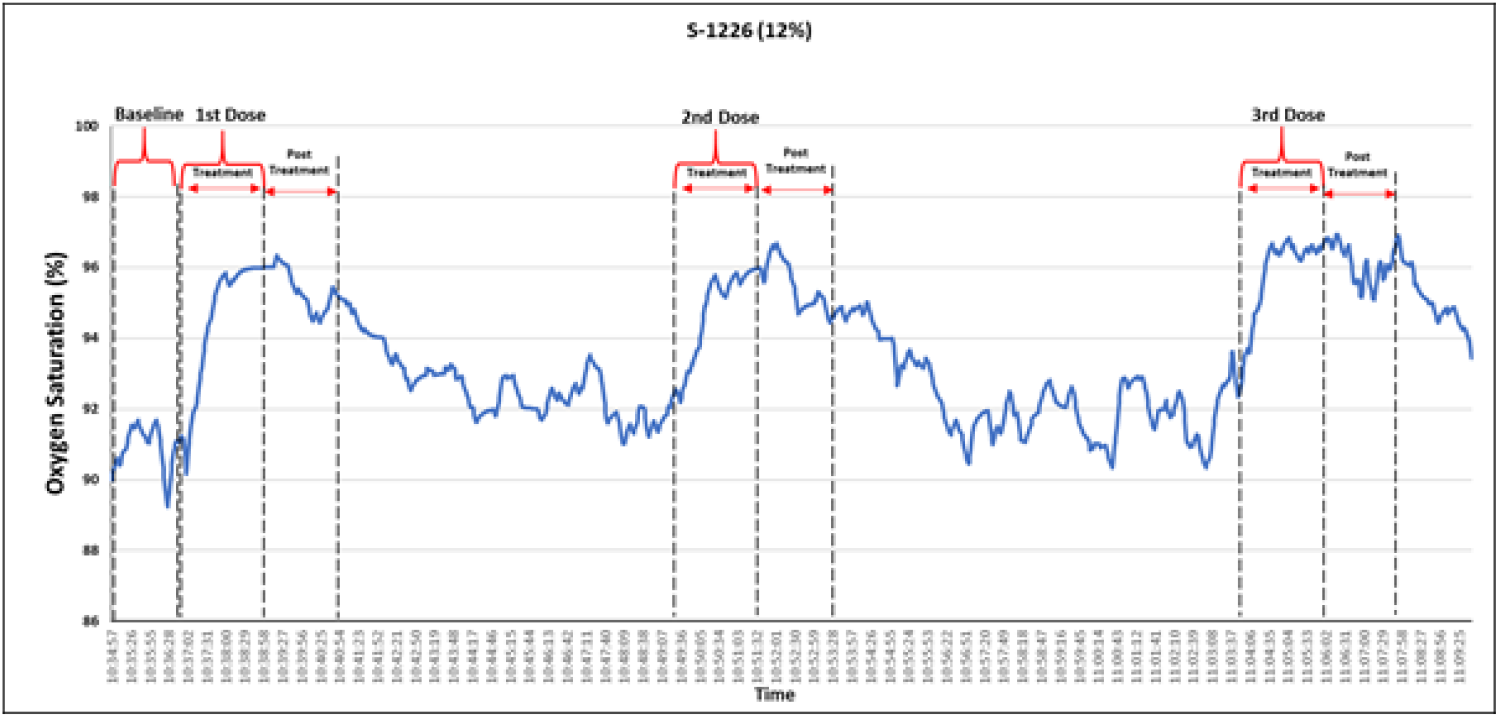
Smoothed Oxygen Saturation (SpO2) graph for CFT01 given S-1226 (12%). This figure illustrates the oxygen saturation for one treatment of a CF Subject (CFT01) during the dose escalation week of the clinical trial. The vertical broken lines show the SpO2 at baseline, during the 2-minute treatment period with S-1226 and for 2-minutes after treatment. This was a typical response for all subjects.

To assess differences in ETCO_2_ and SpO_2_ values at baseline and following treatment a Mean Difference (MD) was calculated from the absolute differences at a 95% CI. The exact test was used to evaluate significance at a α= 0.05.

#### Efficacy evaluations

To analyse for changes in lung function, mean changes in FEV_1_ from baseline after treatment were calculated and expressed as percent change from baseline and a two-tailed paired t-test performed. A p-value of less than 0.05 was considered significant for all analyses. Absolute changes in Percent Predicted FEV_1_ (ppFEV_1_) from baseline following treatment were recorded. Other exploratory efficacy end points were analysed in the same manner. These included Forced Expiratory Flow at 25-75% (FEF_25-75%_), Forced Vital Capacity (FVC) and Lung Clearance Index (LCI). All were reported as relative % change from baseline. To analyse for significant changes in respiratory symptom scores on the Cystic Fibrosis Respiratory Domain questionnaire (CFQ-R), a mean change in the self-reported scores after treatments was calculated from baseline. These scores were evaluated for a minimally clinical important difference (MCID) previously defined as 4.0 for stable patients and 8.5 for CF subjects that experience an exacerbation (Quittner et al., 2009). Mean changes in sputum weight were limited to three individuals and their results are shown descriptively. All parameters are reported as (Mean ± Standard Deviation).

## RESULTS

All subjects were screened and assigned a participant ID (CFT01-05,07). All 6 participants completed the study and follow up visits (**Figure 2**).

**Figure 2:**
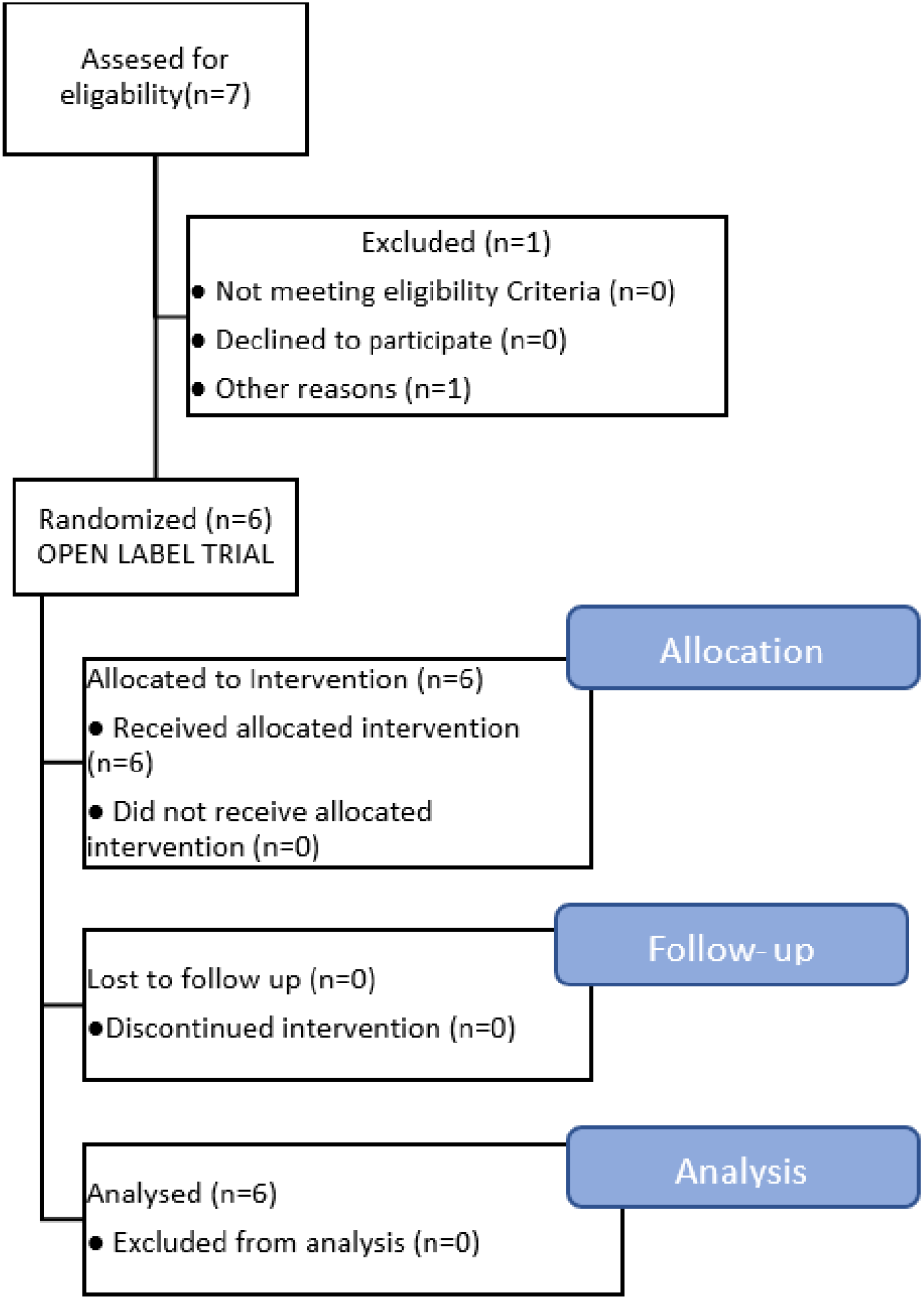
Participant flow diagram following CONSORT Guidelines.

Their ages ranged from 16-31; demographic characteristics and baseline data is summarized in **Table 2**.

**Table 2:**
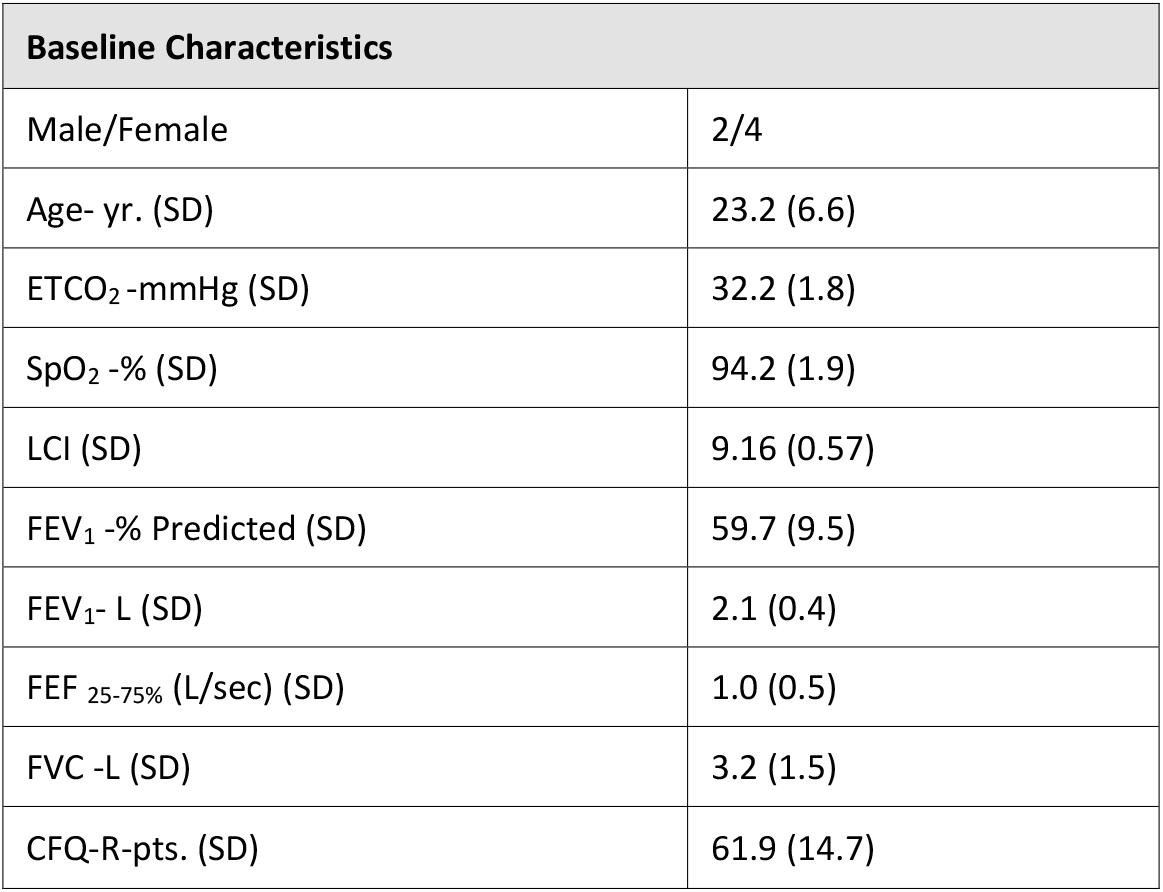
Baseline Characteristics for all subjects (n=6). Values are represented at Mean (Standard Deviation)

### Adverse Events

No Serious Adverse Events (SAEs) were documented at any dose level of S-1226 (4,8,12% CO_2_) for any individual treatments (up to 48 doses administered) that subjects received. Adverse events (AEs) are documented in **Table 3 and Supplemental Table B1**. There were 5 reported AEs in 3 subjects that were considered probably related to the study drug, these included dizziness (N=1), headache (N=1), somnolence(N=1), and flushing (N=1). All were considered mild and rapidly resolved spontaneously without sequelae. These four AEs were similar to those reported in our previous Phase I and II clinical trials (Green et al 2016, Swystun et al, 2018) and attributable to the known effects of inhaled carbon dioxide (Brackett, 1965; Maresh et al, 1997). None recurred on repeat administration of S-1226.

**Table 3.**
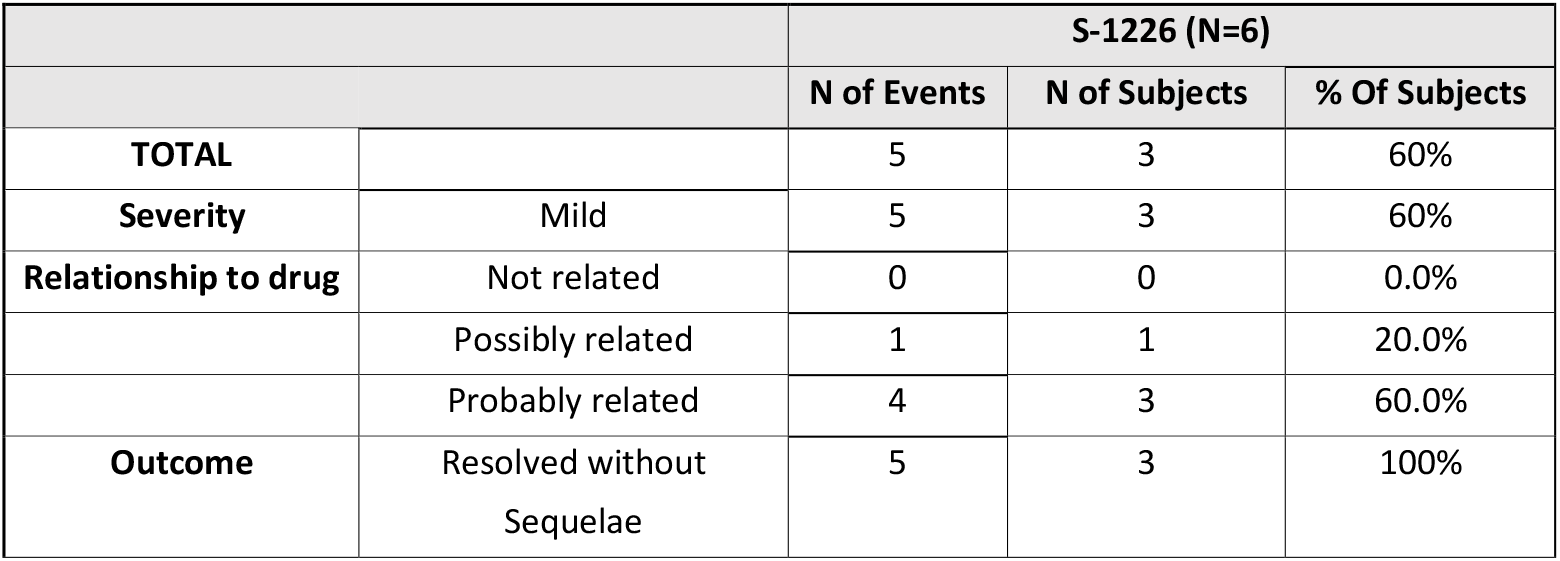
Adverse Events (AEs) by Severity, Relation to Drug, and Outcome in the preliminary Phase IIa Trial Results in Patients with Cystic Fibrosis (CF).

One subject (CFT01) developed a CF related exacerbation at the end of the dose ranging week. The Data safety and monitoring committee determined the exacerbation to be most likely due to infection acquired from a close contact; however, a medication related exacerbation could not be definitively ruled out. The subject’s progress to later stages of the trial was placed on hold and resumed once the exacerbation had resolved. The subject completed subsequent treatments with no further AEs. This AE was included in **Table 3** below as possibly related.

### End Tidal Carbon Dioxide (ETCO_2_) and Oxygen Saturation (SpO_2_)

ETCO_2_ values remained within all subject’s normal range with no significant increases following treatment with S-1226 at 4% CO_2_ (30.1 ±2.2 mmHg), 8% CO_2_ (30.3 ±2.3mmHg) and 12% CO_2_ (31.1 ±2.2mmHg). Notably, a small decrease in ETCO_2_ was observed post treatment when compared against the subject’s values 2 minutes before treatment (Mean Difference (MD) =-1.34; 95% CI= -2.0 to -0.7) These differences were statistically significant, (p value= 0.01). Data for an individual subject is shown in **Supplemental Figure C1 and** summarised for all subjects in **Supplemental Table C1**.

SpO_2_ values at baseline were at or below the normal range, (SpO_2_≤94%), for three of the six subjects. Treatment with S-1226 resulted in a return to a normal of SpO_2_ (>94%) or an increase in SpO_2_ over baseline values for all subjects. The increase in SpO_2_ was rapid and relatively short-term (30 seconds to-20 mins) during and following treatment. For an example see **Figure1**. The improvement was statistically significant and sustained at the 10-14 day follow up when compared to baseline SpO_2_ values (Mean Difference in %SpO_2_ = 1.8; 95% CI= 0.4 to 2.3; p value=0.05). **Figure 3** illustrates this relationship for all subjects.

**Figure 3:**
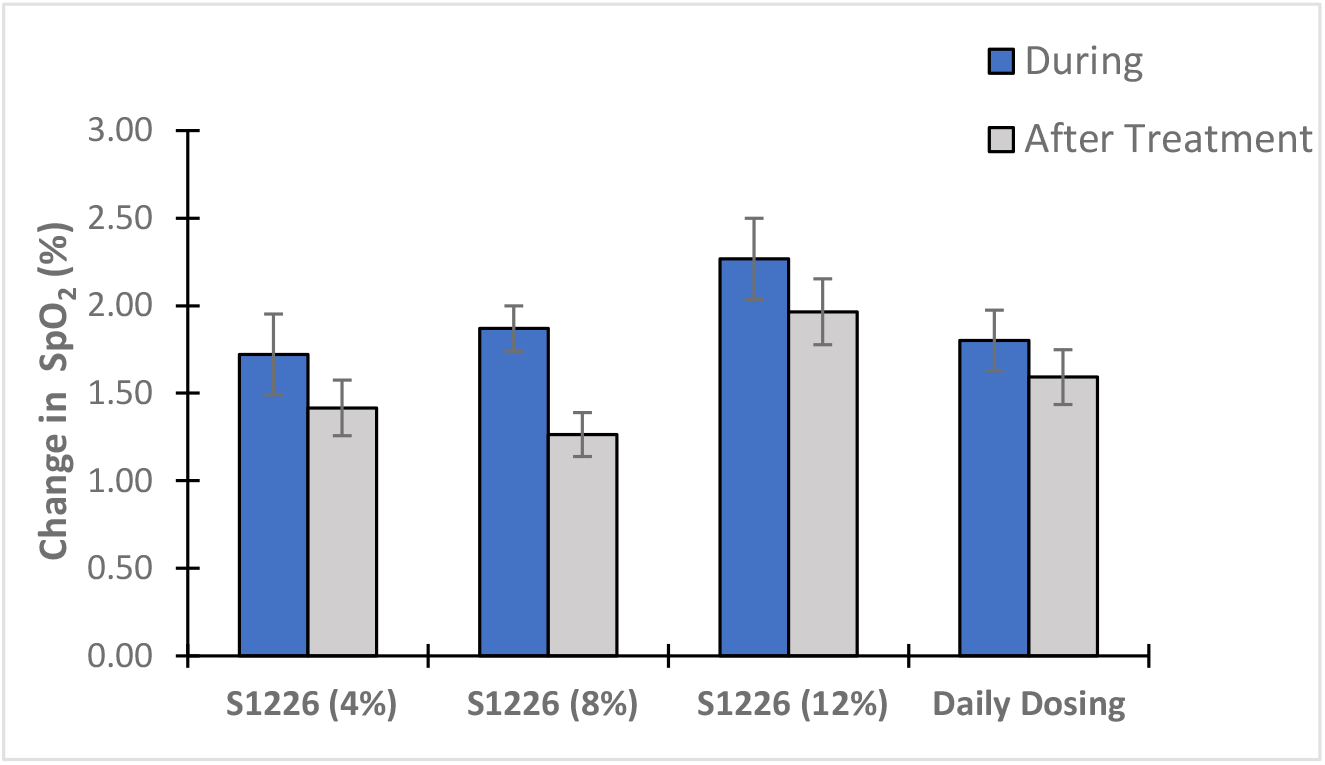
Absolute Change in % SpO_2_ from baseline during (0-2mins) and immediately following treatment (2-4mins). for all subjects (n=6). All subjects showed increases in SpO_2_ above baseline for all three concentrations of S-1226, (4, 8 and 12%). Subjects also showed a dose response relationship following treatment with S-1226. Error bars represent ± Standard Error (SE.

Additionally, there was a dose response effect to S-1226 with increasing concentrations from 4% CO_2_ (Mean Difference in %SpO_2_ = 1.7 ± SD = 1.4); 8% CO_2_(Mean Difference in %SpO_2_ = 1.9 ± SD= 0.8); to 12% CO_2_(Mean Difference in %SpO_2_ = 2.3 ± SD= 1.4) when compared to %SpO_2_ at baseline. There was no evidence of tachyphylaxis (tolerance). In fact, there appeared to be a general upward trend in SpO_2_ with each successive treatment, suggesting a cumulative beneficial effect (**Supplemental Figure D1, and Table D1**).

### Pulmonary Function Tests (PFT) and Lung Clearance Index (LCI)

On average, percent predicted FEV_1_ declined by -0.6%(SD ± 5.1%) by the final day of treatment (Mean= 59.1% ± SD= 11.2%) when compared to baseline values (**Table 2**)). An evaluation of %change in spirometry (FVC, FEF_25-75%,_FEV_1_), showed marked inter-individual variation. (See **Supplemental Figures E1-6)**. On average absolute FVC, FEF_25-75%,_ FEV_1_ declined during the dose escalation week, thereafter they remained unchanged from baseline **(Figure 4)**.

**Figure 4:**
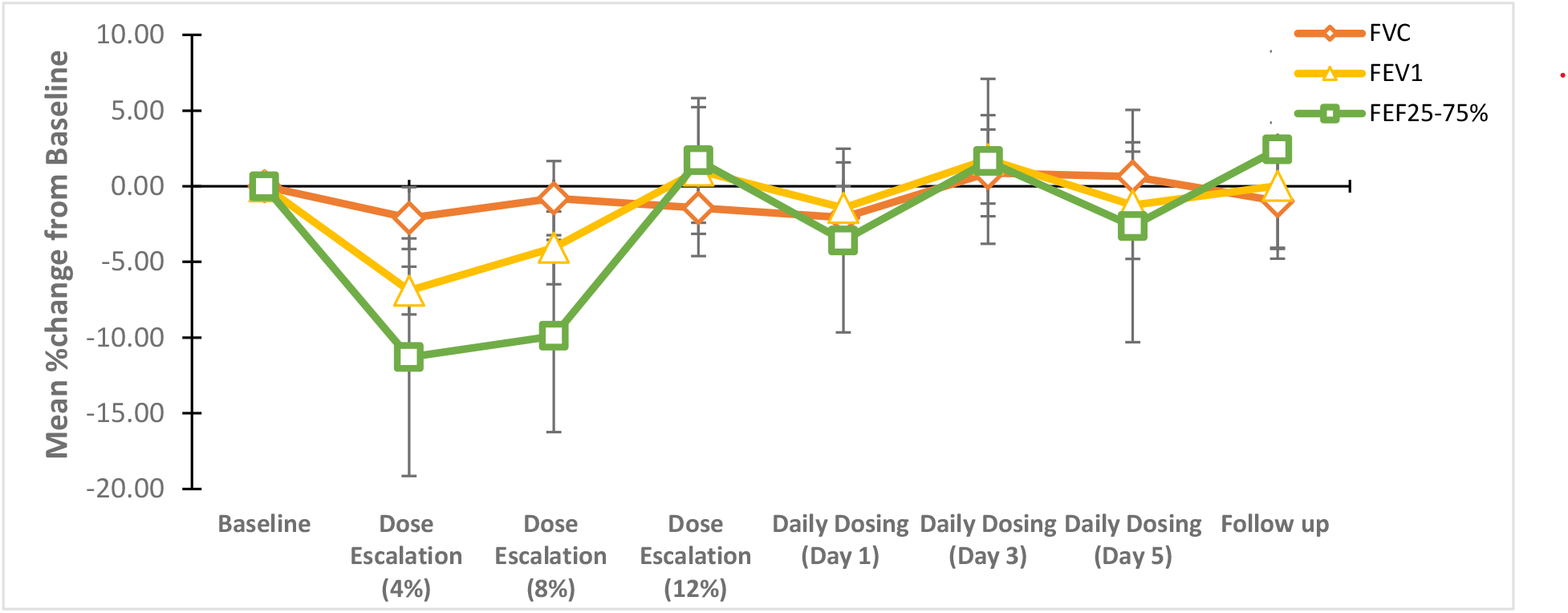
Spirometry results for all subjects (N=6). Values represent the average %change in FVC, FEV_1_ and FEF_25-75%_ absolute values from baseline ±SE (error bars). The first week of the study period included Dose Escalation 4,8 and 12%. The second week of treatment includes Daily Dosing Day 1-5. Follow up was conducted 10-14 days after the final treatment.

Evaluation of %change in LCI provided a consistent measure of a S-1226 treatment effect with an average decrease (improvement) of 6.9% (SD= ±0.2; p =0.4) by the final day treatment. This improvement was sustained at follow up (10-14 days after final treatment) with an overall decline of 8.4% (± SD= 0.08; p = 0.08) compared to baseline (**Figure 5**). Individual subject data for LCI is shown in **Supplemental Figures E1-D to E6-D**.

**Figure 5:**
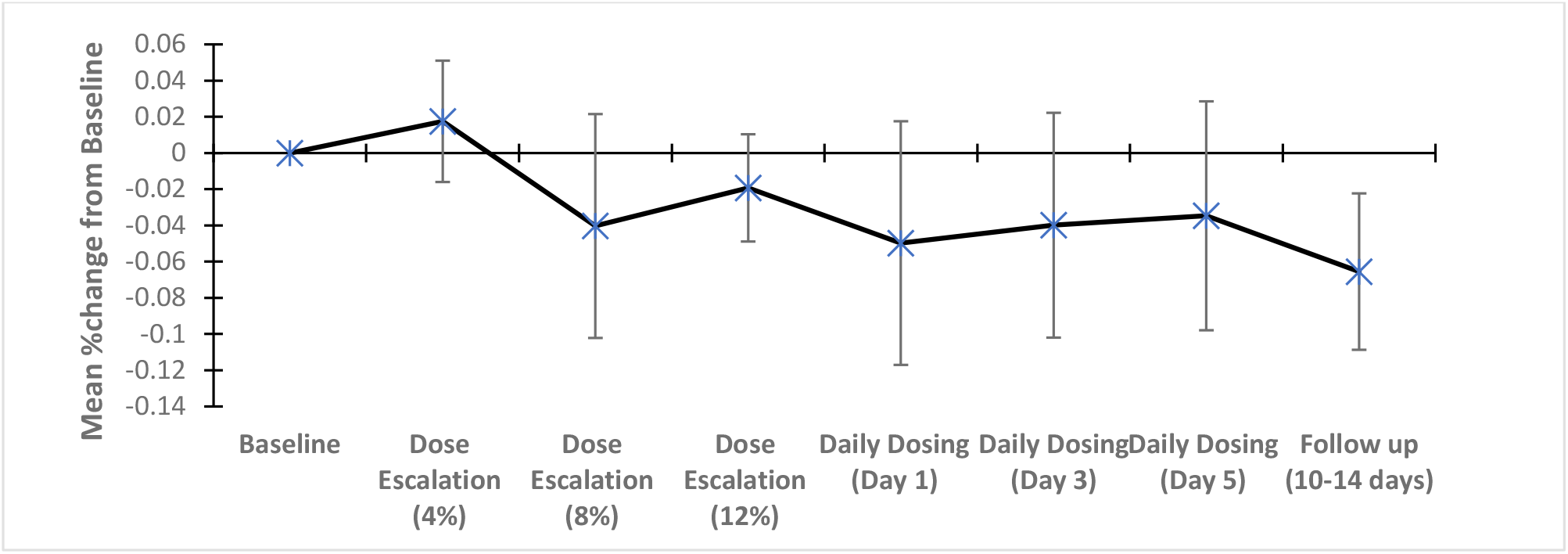
Graph showing LCI %change from baseline for all subjects (N=6). Values represent the mean change (%) in LCI from baseline ±SE (error bars). The first week of the study period included Dose Escalation with 4,8 and 12% CO_2_). The second week of treatment includes Daily Dosing Days 1-5. Follow up was conducted 10-14 days after the final treatment. A lower absolute value indicates a positive treatment effect. **Note:** LCI graphs for individual CF subjects in supplementary Figures E-D are represented as % improvement in LCI values and are thus positive.

### Sputum collection

Sputum samples were successfully collected for CFT02, CFT04, CFT07. All three experienced an increase in sputum weight (g) within the first week of treatment by 14% (CFT02); 64% (CFT04) and 29% (CFT07). CFT04 was the most consistent subject in providing samples over the course of the study. Sputum weight for CFT04 gradually increased from baseline (during the dose escalation period, peaked at daily dosing day 1 (30% increase from baseline) and declined towards baseline for the remainder of the period **(Supplemental Figure F1)**. An example of the expectorated sputum is shown in **(Supplemental Figure F2**). For subjects that produced reliable samples of mucus, the expectorated mucus following S-1226 treatment was less adhesive to the walls of the collection tubes compared to baseline sputum samples. **Figure 6** illustrates this for CFT02.

**Figure 6:**
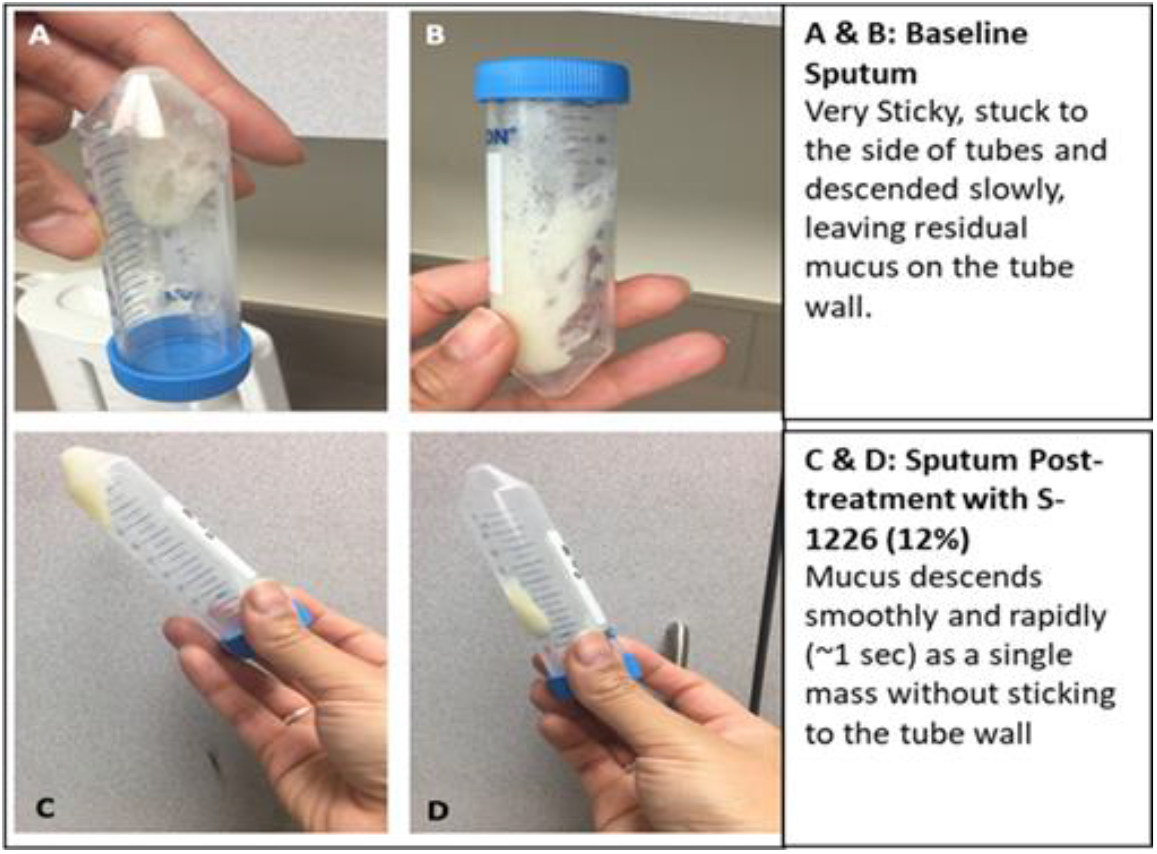
CFT02 Comparison of sputum tenacity (viscosity/adhesiveness). Expectorated mucus sample before (A &B) and after (C & D) treatment with S-1226 (12%).

Sputum volumes tended to increase during the dose ranging period and declined during the daily dosing week. An example of this is shown for subject CFT07 in **Supplemental figure F3**. This subject also received an additional month of home treatment with S-1226.

### Health related Quality of Life assessment (HRQoL): CFQ-R

Health related Quality of Life was assessed by the CF Questionnaire Revised -Respiratory Domain (CFQ-R). On average, subjects reported a 9.5-point increase (SD, ±6.7 points) in their CFQ-R scores, suggesting an overall improvement in health-related quality of life. Of important note, was the improvement in the suppression of irritant cough with continued use of S-1226. All subjects self-reported coughing less frequently, results reflected on the questionnaire. Individually, three subjects (CFT01, CFT02, CFT07) achieved a MCID score over 4.0 points by the end of treatment. CFT04 came close to an MCID with an average increase of 3.4 points. Scores for one subject were excluded from MCID analysis to eliminate the potential for floor or ceiling effects (baseline score was > 90).

### Home Extension Study

One subject, CFT07, successfully completed the 4-week home extension study. Subject reported being able to easily use drug delivery equipment while at home. Similar trends in increase of SpO_2_ and a slight decline ETCO_2_ values were observed as described above **(Supplemental Tables D1 and C1 respectively)**. FVC, FEF_25-75%,_ FEV_1_ values were variable during treatments with no discernible change from baseline **(Supplemental Figure E6)**. However, percent change in LCI values successively declined (improved) by 0.69 units or 7.7% by week 2 and 0.93 units or 10.4% by week 4 when compared to LCI values from the prior week (Daily dosing day 5) suggesting a cumulative benefit to continued use of S-1226 for an additional 4 weeks. For this patient, sputum weight (g) peaked during the first week of treatment but began to decline back to baseline during the home extension treatment **(Supplemental Figure F3**).

## DISCUSSION

The primary aim of this study was to evaluate the safety and tolerability of S-1226 with ascending doses of carbon dioxide (4%, 8%, and 12% CO_2_) in subjects with moderate Cystic Fibrosis Lung disease.

All 6 CF subjects tolerated S-1226 to the maximum dose of 12% CO_2_/3mL nebulized perflubron with up to three doses delivered twice a day. All subjects reported that the drug was easy to administer, non-irritant and eased breathing immediately, several reported marked relief from nasal congestion. The AEs reported were minor and are well known effects of inhaled CO_2_ for tolerance/toxicity (Green *et al*., 2016, Guais et al 2011). All were noted on first administration of S-1226 (4%), resolved rapidly, and without recurrence upon further treatments at same or higher doses. ETCO_2_ was used to further evaluate safety. Importantly, all concentrations of S-1226 were well tolerated and values of ETCO_2_ (pre and post treatment) were within the normal range for all subjects. This finding implies that there were no systemic effects of the inhaled CO_2._The secondary goal of this trial was to evaluate efficacy of S-1226 in CF subjects. Demonstrating efficacy for CF drugs in short duration clinical trials is challenging. The pulmonary disease exhibits many different pathophysiological aspects including mucus hyper-viscosity, impaired mucus clearance, chronic bronchiolitis with surfactant impairment with secondary infection involving many different organisms and culminating in bronchiectasis, which is patchy in distribution. All of these require very different outcome measures to assess. For this reason, we looked at a range of outcome measures in this pilot study.

Results from the participants blood oxygen saturation (SpO_2_) showed an increase in all subjects during and after each treatment. This increase, though relatively short term, was still evident at the final day of treatment where all subjects on average improved by 2% from their baseline %SpO_2_ **(Figure 3)**. Additionally, SpO_2_ data showed no evidence for tachyphylaxis (tolerance) over three consecutive treatments, twice daily, for 14 days. These beneficial effects were most likely due to the extrinsic CO_2_ as there was a trend for a dose response relationship from 4 to 12% CO_2._ The mechanism for this improvement in blood oxygenation is not entirely clear. None of the subjects received supplementary oxygen and the oxygen concentration in the medical gas tanks was maintained at 21%. A probable explanation are the known effects of extrinsic carbon dioxide as a potent small airway bronchodilator with ability to improve ventilation/perfusion (V_A_/Q) matching (Brogan *et al*., 2004). Other benefits of extrinsic CO_2_ include improved collateral ventilation (Traystman *et al*., 1978), maintaining a low surface tension of alveolar surfactant (Wildeboer-Venema *et al*., 1984), and facilitating local changes in lung compliance that favour improved 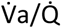 matching, (Emery et al, 2007). None of these effects are described for high intrinsic CO_2_(hypercarbia).

Spirometry (FVC, FEF_25-75%_ and FEV_1_) proved the least useful of the pulmonary function tests in assessing the effectiveness of S-1226 in this short-term study. Spirometry, though relatively straightforward and simple to perform is highly effort-dependent and requires active participation from patients to capture accurate and replicable results (Stanojevic & Ratjen, 2016). Some factors that have been found to limit the validity of FEV_1_ results in CF trials include concomitant drug effects, variability within the CF population, spirometry performance, cough and mucus movement in the airways (Casier *et al*., 2013; Stanojevic & Ratjen, 2016; Rubin, 2007, Vilozni *et al*., 2014). The latter effect may have been operative in this study as mucus in transit through the airways would be expected to limit airflow. This effect has been observed in other studies of expectorants in CF, including clinical trials of hypertonic saline (Robinson et al, 1997) and mannitol (Robinson et al, 1999). The small initial declines in FEV_1_ in these studies, as in ours, was followed by a return to predicted levels at study end, see **Figure 4**, indicating that increased mucus flow may have contributed to airflow limitation during the first week of treatment. We expect that in longer term studies, as mucous secretions are cleared from the airways, FEV_1_ will improve. One subject in this study (CFT02) produced very little mucus throughout the study but showed a good response in pulmonary function following treatment with S-1226, particularly for tests of small airway function (FEF_25-75%_) (**Supplemental Figures E2-C**. Small airways are the primary site of inflammation in CF (Vilozni *et al*., 2014). This subject also showed improvements in FEV_1_ and Lung Clearance Index (LCI) (**Supplemental Figures E2-B and D**).

A good indirect index of mucus clearance is the LCI, as this measures the homogeneity of ventilation. It also has been shown to be good predictor of mortality and requirement for lung transplantation in CF patients (Kurz et al., 2021). We observed a general improvement in LCI for all subjects, with four subjects showing a consistent %change improvement in LCI over the course of the study **(Supplemental Figures E2-D to E4-D)**. This improvement was sustained at the 10-14 day follow up, suggesting that the effects of daily S-1226 use extends 10-14 days post treatment. Overall, LCI appears to be a better outcome measure for early CF lung disease than FEV_1_ as it provides an index of the homogeneity of ventilation over the whole lung, whereas FEV_1_ is primarily a measure of large airway flow that may be compromised by mucous plugs (Gustafsson *et al*., 2008). Measures of statistical significance for improvement in ppFEV1 and/or LCI may not be useful for this study based on the small number of CF subjects enrolled (n=6). However, these results will inform future sample size calculations in later trials.

Mucus clearance is an important parameter for measuring efficacy of CF drugs. Three subjects were diligent in collecting their mucus. For these three subjects, sputum percent weight greatly increased from baseline during the dose escalation week. One patient (CFT04) noted that their mucus during treatments was different than normal, being more “solid and clumpy” and that it “looks like what [they] get after receiving antibiotics during an exacerbation” **(Supplemental Figure F2)**. This subject had severe lung disease with a pre-treatment FEV_1_ of 43% of predicted.

Clinical trials face challenges in standardizing data collection when it comes to measuring sputum volume/weight as its variability is affected by many factors including unintentional swallowing, saliva, and variability in cough that affects sputum mobility. It is also affected by demographic and social factors such as age, sex, and cultural background, where sputum collection may be perceived as inappropriate (Rubin, 2007). Sputum production is variable not only from one day to another but also throughout the day, with greater volumes recorded in the mornings. Additionally, it is difficult to distinguish whether an increase in sputum collection is due to increased clearance or production. For this reason, various other measures of mucus clearance have been proposed. These include chest radiography, local tracer, and whole lung tracer imaging, although these methods are ineffective for evaluating changes over short time periods (Rubin, 2007). Future trials with S-1226 may explore these alternatives.

Markers of improvements in health-related quality of Life are important in investigational studies. The Cystic Fibrosis Questionnaire-Revised (CFQ-R) was developed as one such measure to assess symptoms and quality of life specific to CF patients. Four subjects showed improvements in their CFQ-R points, three of whom achieved a clinically significant change assessed by the CFQ-R.

S-1226 appears to be better tolerated compared to hypertonic saline, a respiratory tract irritant. An unexpected finding was that S-1226 completely suppressed irritant cough during and immediately after treatments. We attribute this effect to both the perflubron and CO_2_ components. Perflubron is a surfactant. Surfactants are thought to mask irritant nerve receptors that control airway smooth muscle tone (Koetzler et al., 2006; Hohlfield et al., 1997)) and cough (Hills et al., 2000). Surfactants have been used clinically to suppress irritant cough in patients with tuberculosis (Lovacheva et al., 2006). Extrinsic CO_2_ has been shown to suppress irritant cough in patients with pulmonary tuberculosis (Banyai et al., 1944; Banyai, 1947). In addition, most subjects reported increased ease of breathing during and post treatment, increased sputum volume and reduced nasal congestion during and after treatments. These subjective outcomes will be evaluated in objective manner in future studies.

## CONCLUSIONS

Overall, this preliminary analysis of 6 CF subjects indicates that S-1226 is safe and efficacious in treating common symptoms of CF lung disease. Together these findings provide preliminary evidence and reassurance that clinic and home use trials of S-1226 will be feasible and can be safely studied. Plans to progress the use of S-1226 in the home environment and to extend it to include non-CF patients with bronchiectasis are in place. This latter group is arguably in greater need of a drug that has the potential of S-1226 for treating their disease.

## Data Availability

This trial has been registered at clinical trials.gov NCT03903913. Some data generated or analyzed during this study is included in the Supplemental materials to support the findings of this study. Additional data and study protocol are available upon reasonable request with the permission of SolAeroMed Inc. Please be aware that some restrictions apply to the availability of these data since they were used under license for the current study and are not publicly available.

https://www.solaeromed.com/

## ACKNOWLEDGMENTS AND FUNDING

We would like to thank Alberta Lung Function for allowing us to conduct this trial at their site.

This clinical trial was funded in part by SolAeroMed Inc. SolAeroMed Inc. also provided the drug and delivery systems.

## AVAILABILITY OF DATA AND MATERIALS

This trial has been registered at clinicaltrials.gov NCT03903913. Some data generated or analysed during this study is included in the **Supplemental** materials to support the findings of this study. Additional data and study protocol are available upon reasonable request with the permission of SolAeroMed Inc. Please be aware that some restrictions apply to the availability of these data since they were used under license for the current study and are not publicly available.

## REFERENCES

Al-Saiedy M, Nelson E, El-Mays T, Amrein M, Green F. Perflubron enhances mucin plug clearance in vitro in the presence of natural surfactant, 2013. Eur Respiratory Soc poster

Albers GM, Tomkiewicz RP, May MK, Ramirez OE, Rubin BK. Ring distraction technique for measuring surface tension of sputum: relationship to sputum clearability. J Appl Physiol 1996;81(6):2690–2695

Anzueto A1, Peters JI, Tobin MJ, de los Santos R, Seidenfeld JJ, Moore G, Cox WJ, Coalson JJ. Effects of prolonged controlled mechanical ventilation on diaphragmatic function in healthy adult baboons. Crit Care Med. 1997 Jul;25(7):1187–90.

Banyai AL, Cadden AV. Carbon dioxide by inhalation in the management of cough: With observations on its effects upon respiration in pulmonary tuberculosis. British Journal of Tuberculosis and Diseases of the Chest. 1944 Oct 1;38(4):111–6.

Banyai AL. Fifteen years’ experience with carbon dioxide in the management of cough. Diseases of the Chest. 1947 Jan 1;13(1):1–9.

Brackett CN, Cohen JJ, Schwartz WB. Carbon dioxide titration curve of normal man. Effects of increasing degrees of acute hypercapnia on acid-base equilibrium. NEJM. Jan 1965; 272 (1): 6–12.

Brewington JJ, Backstrom J, Feldman A, Kramer EL, Moncivaiz JD, Ostmann AJ, et al. Chronic beta2AR stimulation limits CFTR activation in human airway epithelia. JCI Insight. 2018;3(4).

Brogan TV, Robertson HT, Lamm WJ, Souders JE, Swenson ER. Carbon dioxide added late in inspiration reduces ventilation-perfusion heterogeneity without causing respiratory acidosis. J Appl Physiol 2004; 96(5): 1894–1898.

Casier A, Goubert L, Vervoort T, Theunis M, Huse D, De Baets F, Matthys D, Crombez G. Spirometry-related pain and distress in adolescents and young adults with cystic fibrosis: The role of acceptance. Pain Research and Management. 2013 Nov 1;18(6):286–92.

Choudhury P, El Mays T, Snibson K, Wilson R, Leigh R, Dennis J, et al. Investigations of mechanisms of carbon dioxide-induced bronchial smooth muscle relaxation. Am J Respir Crit Care Med. 2012;185:A2848.

Coakley RJ, Taggart C, Greene C, McElvaney NG, O’Neill SJ. Ambient pCO2 modulates intracellular pH, intracellular oxidant generation, and interleukin-8 secretion in human neutrophils. Journal of leukocyte biology. 2002 Apr;71(4):603–10.

Croce H Jr, Melton SM, Moore M, Trenthem LL. Partial liquid ventilation decreases the inflammatory response in the alveolar environment of trauma patients. J Trauma 1998; 45(2):273–280.

Cystic Fibrosis Canada. (2018). Canadian Cystic Fibrosis Registry. Annual Data Report 2018. www.cysticfibrosis.ca.:https://www.cysticfibrosis.ca/uploads/RegistryReport2018/2018RegistryAnnualDataReport.pdf

Dagenais RV, Su VC, Quon BS. Real-world safety of CFTR modulators in the treatment of cystic fibrosis: A systematic review. Journal of Clinical Medicine. 2021 Jan;10(1):23.

De Sanctis GT, Tomkiewicz RP, Rubin BK, Schurch S, King M. Exogenous surfactant enhances mucociliary clearance in the anaesthetized dog. European Respiratory Journal. 1994 Sep 1;7(9):1616–21.

Dorrington KL, Balanos GM, Talbot NP, Robbins PA. Extent to which pulmonary vascular responses to PCO2 and PO2 play a functional role within the healthy human lung. J Appl Physiol 2010; 108(5): 1084–1096.

El-Betany AM, Behiry EM, Gumbleton M, Harding KG. Humidified warmed CO2 treatment therapy strategies can save lives with mitigation and suppression of SARS-CoV-2 infection: an evidence review. Frontiers in Medicine. 2020 Dec 11; 7:982.

El Mays TY, Choudhury P, Leigh R, Koumoundouros E, Van der Velden J, Shrestha G, et al. Nebulized PFOB and carbon dioxide rapidly dilate constricted airways in an ovine model of allergic asthma. Respir Res. 2014; 15:98.

El Mays TY, Saifeddine M, Choudhury P, Hollenberg MD, Green FH. Carbon dioxide enhances substance P-induced epithelium-dependent bronchial smooth muscle relaxation in Sprague–Dawley rats. Can J Physiol Pharmacol. 2011, 89(7):513–520.

Fahy JV and Dickey BF. Airway mucus function and dysfunction. N Engl J Med. 2010;363(23):2233–47.

Fisher HK, Hansen TA. Site of action of inhaled 6 per cent carbon dioxide in the lungs of asthmatic subjects before and after exercise. Am Rev Respir Dis. 1976;114(5):861–70.

Fleischmann E, Herbst F, Kugener A, Kabon B, Niedermayr M, Sessler DI, Kurz A. Mild hypercapnia increases subcutaneous and colonic oxygen tension in patients given 80% inspired oxygen during abdominal surgery. Anesthesiology 2006; 104(5): 944–949.

Flume PA, Van Devanter DR. State of progress in treating cystic fibrosis respiratory disease. BMC Med. 2012; 10:88.

Gerber F, Krafft MP, Vandamme TF, Goldmann M, Fontaine P. Potential use of fluorocarbons in lung surfactant therapy. Artificial cells, blood substitutes, and biotechnology. 2007 Jan 1;35(2):211–20.

Gates KL, Howell HA, Nair A, Vohwinkel CU, Welch LC, Beitel GJ, Hauser AR, Sznajder JI, Sporn PH. Hypercapnia impairs lung neutrophil function and increases mortality in murine pseudomonas pneumonia. American journal of respiratory cell and molecular biology. 2013 Nov;49(5):821–8.

Gerber F, Krafft MP, Vandamme TF, Goldmann M, Fontaine P. Potential use of fluorocarbons in lung surfactant therapy. Artif Cells Blood Substit Immobil Biotechnol. 2007;35(2):211–20.

Green FH, Leigh R, Fadayomi M, Lalli G, Chiu A, Shrestha G, ElShahat SG, Nelson DE, El Mays TY, Pieron CA, Dennis JH. A phase I, placebo-controlled, randomized, double-blind, single ascending dose-ranging study to evaluate the safety and tolerability of a novel biophysical bronchodilator (S-1226) administered by nebulization in healthy volunteers. Trials. 2016 Dec;17(1):361.

Griese M, Birrer P, Demirsoy A. Pulmonary surfactant in cystic fibrosis. Eur Respir J. 1997;10(9):1983–8.

Griese M, Essl R, Schmidt R, Ballmann M, Paul K, Rietschel E, et al. Sequential analysis of surfactant, lung function and inflammation in cystic fibrosis patients. Respir Res. 2005; 6:133.

Guais A, Brand G, Jacquot L, Karrer M, Dukan S, Grévillot G, et al. Toxicity of carbon dioxide: a review. Chem Res Toxicol. (2011) 24:2061–70. doi: 10.1021/tx200220r

Gunasekara L, Al-Saiedy M, Green F, Pratt R, Bjornson C, Yang A, Schoel WM, Mitchell I, Brindle M, Montgomery M, Keys E. Pulmonary surfactant dysfunction in pediatric cystic fibrosis: Mechanisms and reversal with a lipid-sequestering drug. Journal of Cystic Fibrosis. 2017 Sep 1;16(5):565–72.

Gustafsson PM, De Jong PA, Tiddens HA, Lindblad A. Multiple-breath inert gas washout and spirometry versus structural lung disease in cystic fibrosis. Thorax. 2008 Feb 1;63(2):129–34.

Habib AR, Kajbafzadeh M, Desai S, Yang CL, Skolnik K, Quon BS. A systematic review of the clinical efficacy and safety of CFTR modulators in cystic fibrosis. Scientific reports. 2019 May 10;9(1):1–9.

Hills BA, Chen Y. Suppression of neural activity of bronchial irritant receptors by surface-active phospholipid in comparison with topical drugs commonly prescribed for asthma. Clinical & Experimental Allergy. 2000 Sep;30(9):1266–74.

Hohlfeld J, Hoymann HG, Molthan J, Fabel H, Heinrich U. Aerosolized surfactant inhibits acetylcholine-induced airway obstruction in rats. European Respiratory Journal. 1997 Oct 1;10(10):2198–203.

Hoiby N, Ciofu O, Bjarnsholt T. Pseudomonas aeruginosa biofilms in cystic fibrosis. Future Microbiol. 2010;5(11):1663–74.

Jackson, L., DePas, W., Morris, A. J., Guttman, K., Yau, Y. C. W., & Waters, V. (2020). Visualization of Pseudomonas aeruginosa within the Sputum of Cystic Fibrosis Patients. Journal of Visualized Experiments: JoVE, 161. https://doi.org/10.3791/61631

Koetzler R, Saifeddine M, Yu Z, Schurch FS, Hollenberg MD, Green FH. Surfactant as an airway smooth muscle relaxant. American journal of respiratory cell and molecular biology. 2006 May;34(5):609–15.

Kurz, J. M., Ramsey, K. A., Rodriguez, R., Spycher, B., Biner, R. F., Latzin, P., & Singer, F. (2021). Association of lung clearance index with survival in individuals with cystic fibrosis. European respiratory journal.

Lehmler HJ, Bummer PM, Jay M. Liquid ventilation-A new way to deliver drugs to diseased lung? Chemtech. 1999; 29(10):7–12.

Lovacheva, O., Erokhin, W., Chernichenko, N., Evgushchenko, G., Lepekha, L., & Rozenberg, O. (2006). esults of use of surfactant in complex therapy of patients with destructive pulmonary tuberculosis. Problemy Tuberkuleza I Boleznei Legkikh, (10), 12–17.

Lyczak JB, Cannon CL, Pier GB. Lung infections associated with cystic fibrosis. Clin Microbiol Rev. 2002;15(2):194–222.

Maresh CM, Armstrong LE, Kavouras SA, Allen GJ, Casa DJ, Whittlesey M, et al. Physiological and psychological effects associated with high carbon dioxide levels in healthy men. Aviat Space Environ Med. Jan 1997; 68(1):41–5.

Mogayzel, P. J., Naureckas, E. T., Robinson, K. A., Mueller, G., Hadjiliadis, D., Hoag, J. B., Lubsch, L., Hazle, L., Sabadosa, K., & Marshall, B. (2013). Cystic Fibrosis Pulmonary Guidelines. American Journal of Respiratory and Critical Care Medicine, 187(7), 680–689. https://doi.org/10.1164/rccm.201207-1160oe

Montgomery ST, Mall MA, Kicic A, Stick SM, Arest CF. Hypoxia and sterile inflammation in cystic fibrosis airways: mechanisms and potential therapies. Eur Respir J. 2017;49(1).

Quittner, A. L., Modi, A. C., Wainwright, C., Otto, K., Kirihara, J., & Montgomery, A. B. (2009). Determination of the Minimal Clinically Important Difference Scores for the Cystic Fibrosis Questionnaire-Revised Respiratory Symptom Scale in Two Populations of Patients With Cystic Fibrosis and Chronic Pseudomonas aeruginosa Airway Infection. Chest, 135(6), 1610–1618. https://doi.org/10.1378/chest.08-1190

Ratjen FA. Cystic fibrosis: pathogenesis and future treatment strategies. Respir Care. 2009;54(5):595–605.

Robinson M, Hemming AL, Regnis JA, Wong AG, Bailey DL, Bautovich GJ, King M, Bye PT. Effect of increasing doses of hypertonic saline on mucociliary clearance in patients with cystic fibrosis. Thorax. 1997 Oct;52(10):900.

Robinson M, Daviskas E, Eberl S, Baker J, Chan HK, Anderson SD, Bye PP. The effect of inhaled mannitol on bronchial mucus clearance in cystic fibrosis patients: a pilot study. European Respiratory Journal. 1999 Sep 1;14(3):678–85.

Rotta AT, Steinhorn DM. Partial liquid ventilation reduces pulmonary neutrophil accumulation in an experimental model of systemic endotoxemia and acute lung injury. Crit Care Med 1998; 26(10):1707–1715.

Rubin, B. K. (2007). Designing clinical trials to evaluate mucus clearance therapy. Respiratory care, 52(10), 1348–1361. http://rc.rcjournal.com/content/52/10/1348.short

Schürch S. Perfluorocarbon-surfactant interactions: biophysical aspects. Am Soc Artif Intern Organs 2006, 52:487.

Sinclair SE, Kregenow DA, Starr I, Schimmel C, Lamm WJ, Hlastala MP, Swenson ER. Therapeutic hypercapnia and ventilation-perfusion matching in acute lung injury: low minute ventilation vs inspired CO2. Chest. 2006 Jul 1;130(1):85–92.

Stanojevic, S., & Ratjen, F. (2016). Physiologic endpoints for clinical studies for cystic fibrosis. Journal of Cystic Fibrosis, 15(4), 416–423. https://doi.org/10.1016/j.jcf.2016.05.014

Strandvik B. Fatty acid metabolism in cystic fibrosis. Prostaglandins Leukot Essent Fatty Acids. 2010;83(3):121–9.

Swystun V, Green FH, Dennis JH, Rampakakis E, Lalli G, Fadayomi M, Chiu A, Shrestha G, El Shahat SG, Nelson DE, El Mays TY. A phase IIa proof-of-concept, placebo-controlled, randomized, double-blind, crossover, single-dose clinical trial of a new class of bronchodilator for acute asthma. Trials. 2018 Dec 1;19(1):321.

Tang SE, Wu SY, Chu SJ, Tzeng YS, Peng CK, Lan CC, Perng WC, Wu CP, Huang KL. Pre-treatment with ten-minute carbon dioxide inhalation prevents lipopolysaccharide-induced lung injury in mice via down-regulation of toll-like receptor 4 expression. International journal of molecular sciences. 2019 Jan;20(24):6293.

Tawfic QA, Kausalya R. Liquid ventilation. Oman Med J 2011, 26(1): 4–9.

Thome UH, Ambalavanan N. Permissive hypercapnia to decrease lung injury in ventilated preterm neonates. Semin Fetal Neonatal Med. 2009;14(1):21–7

Traystman RJ, Terry R, and Menkes HA. Carbon dioxide: a major determinant of collateral ventilation. J Appl Physiol 45: 69–74, 1978

Twort CH, Cameron IR. Effects of P CO2, pH and extracellular calcium on contraction of airway smooth muscle from rats. Respir Physiol 1986; 66(3):259-67

Van den Elshout FJ, Van Herwaarden CL, Folgering HT. Effects of hypercapnia and hypocapnia on respiratory resistance in normal and asthmatic subjects. Thorax. 1991 Jan 1;46(1):28–32.

Vilozni, D., Lavie, M., Ofek, M., Sarouk, I., & Efrati, O. (2014). Cough characteristics and FVC maneuver in cystic fibrosis. Respiratory Care, 59(12), 1912–1917. https://doi.org/10.4187/respcare.03290

Ward, N., Stiller, K., Rowe, H., & Holland, A. E. (2017). The psychometric properties of the Leicester Cough Questionnaire and Respiratory Symptoms in CF tool in cystic fibrosis: A preliminary study. Journal of Cystic Fibrosis, 16(3), 425–432.

Wildeboer-Venema F. Influences of nitrogen, air and alveolar gas upon surface tension of pulmonary surfactant. Respir Physiol 58:1–14, 1984

Wolfson MR, Shaffer TH. Pulmonary applications of perfluorochemical liquids: ventilation and beyond. Paediatr Respir Rev 2005, 6(2):117–127.

