## Supplemental Materials for "A Phase IIa Open Label Study to Evaluate the Safety, Tolerability and Efficacy of S-1226 Administered by Nebulization in Subjects with Cystic Fibrosis Lung Disease"

#### SUPPLEMENTAL A: STUDY SCHEDULE

**Supplemental Figure A1: Study Schedule Overview**

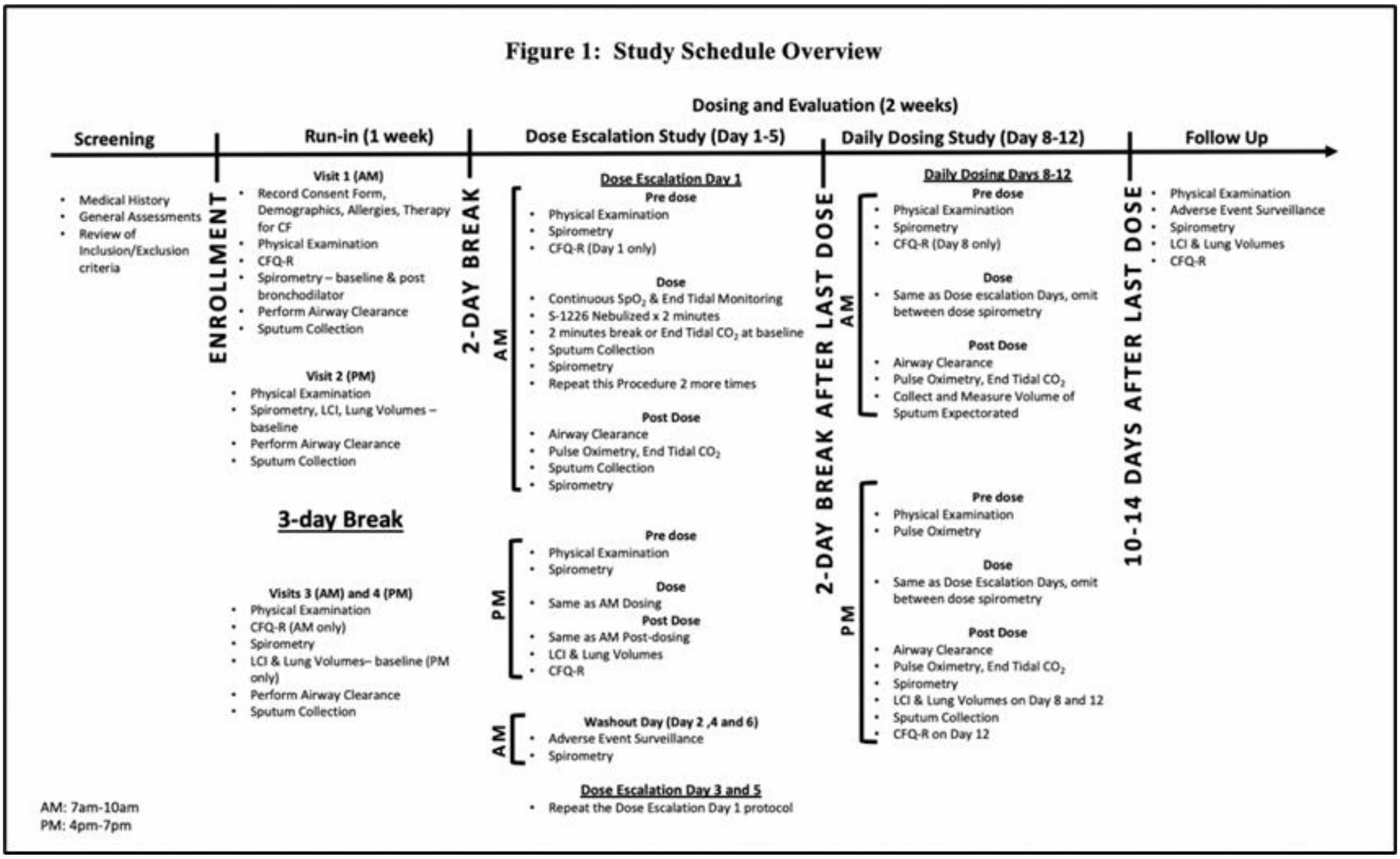

### SUPPLEMENTAL B: ADVERSE EVENTS

**Supplemental Table B1: Incidence of Adverse Events (AEs) by System Organ Class in the preliminary Phase IIa Trial Results in Patients with Cystic Fibrosis (CF).**

| | | S-1226 (N=5) | | |
|---|---|---|---|---|
| **System Organ Class** | **Preferred Term** | **N of Events** | **N of Subjects** | **% of Subjects** |
| **TOTAL AEs** | | 5 | 3 | 60% |
| **Cardiac Disorders** | | 0 | 0 | 0% |
| **Respiratory disorders** | **CF Exacerbation** | 1 | 1 | 20% |
| **General disorders and administration site conditions** | | 0 | 0 | 0% |
| | SUBTOTAL | 1 | 1 | 20% |
| **Nervous system disorders** | **Somnolence**<br>**Dizziness**<br>**Headache** | 1<br>1<br>1 | 1<br>1<br>1 | 20%<br>20%<br>20% |
| | SUBTOTAL | 3 | 3 | 60% |
| **Psychiatric disorders** | **Anxiety** | 0 | 0 | 0% |
| | SUBTOTAL | 0 | 0 | 0% |
| **Vascular disorders** | **Flushing** | 1 | 1 | 20.0% |
| | SUBTOTAL | 1 | 1 | 20.0% |

#### SUPPLEMENTAL C: END TIDAL CARBON DIOXIDE (ETCO$_2$)

ETCO$_2$ (mmHg) was obtained for all subjects at baseline and immediately following all treatments using a nasal canula (Capnostream™ 35 Portable Respiratory Monitor). Values were obtained at each second to generate a times series graph. Exponential smoothing at an appropriate dampening factor ($1 - \alpha$) was carried out on the raw data. This was done to remove high frequency noise within the datasets and improve pattern recognition during analysis.

Figure C1 illustrates the ETCO$_2$ data collected during a typical treatment. The grey columns represent the smoothed ETCO$_2$ while the blue line shows the smoothed SPO$_2$ data for a single dose of S-1226 (8%). The vertical broken lines indicate the two intervals (2 minutes) used in the analyses to include a baseline and a 2- minute post treatment period. Table C1 shows the summary data for all 5 subjects.

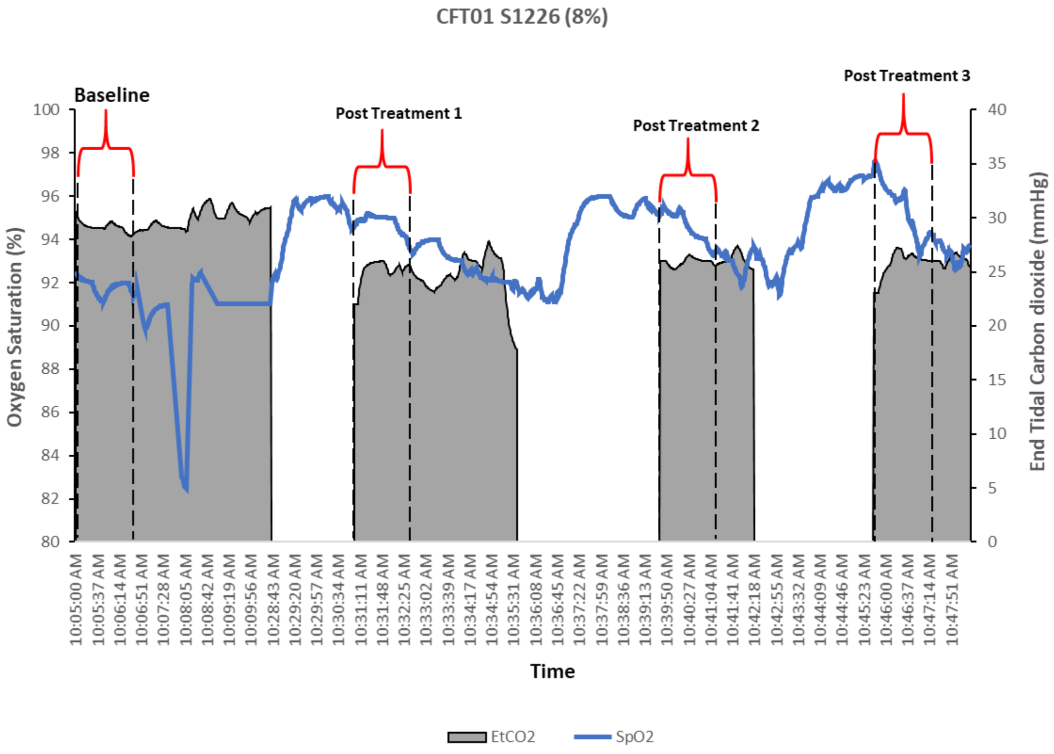


**Supplemental Figure C2**: **Graph showing smoothed SPO$_2$ and ETCO$_2$ for CFT01 at S-1226 (8%)** for one dosing period of three consecutive treatments. End tidal CO$_2$ was averaged for two minutes at baseline and immediately after completion of each treatment, as shown in the vertical dotted bars. Note that ETCO$_2$ is slightly lower post-treatment than pre-treatment. This was a characteristic finding for all doses and subjects.

**Supplemental Table C1: End Tidal Carbon Dioxide ($ETCO_2$) Pre and Post Treatment for all subjects**

| | CFT01 | CFT02 | CFT03** | CFT04 | CFT05 | CFT07 | Averages | SD |
|---|---|---|---|---|---|---|---|---|
| **S1226 (4%)** | | | | | | | | |
| $ETCO_2$ Before | 28.82 | 32.30 | 33.76 | 33.52 | 32.08 | 32.75 | 32.20 | 1.79 |
| $ETCO_2$ Post Treatment | 27.07 | 30.17 | 32.79 | 32.35 | 30.13 | 28.23 | 30.1 | 2.2 |
| **S1226 (8%)** | | | | | | | | |
| $ETCO_2$ Before | 28.93 | 32.89 | 31.97 | 32.46 | 32.60 | 30.63 | 31.58 | 1.52 |
| $ETCO_2$ Post Treatment | 26.12 | 29.98 | 32.37 | 32.01 | 31.32 | 29.92 | 30.3 | 2.3 |
| **S1226 (12%)** | | | | | | | | |
| $ETCO_2$ Before | 29.08 | 33.67 | 33.11 | 31.05 | 33.45 | 32.42 | 32.13 | 1.77 |
| $ETCO_2$ Post Treatment | 26.94 | 31.71 | 33.13 | 30.84 | 32.83 | 31.29 | 31.1 | 2.2 |
| **Daily Dosing** | | | | | | | | |
| $ETCO_2$ Before | 30.17 | 31.84 | 33.90 | 30.69 | 31.61 | 30.31 | 31.42 | 1.39 |
| $ETCO_2$ Post Treatment | 28.40 | 29.60 | 32.49 | 31.24 | 30.13 | 30.16 | 30.3 | 1.4 |
| **Home Extension** | | | | | | | | |
| $ETCO_2$ Before | NOT APPLICABLE | | | | | 31.32 | | |
| $ETCO_2$ Post Treatment | | | | | | 30.24 | | |

#### Supplemental D: Oxygen Saturation

$SpO_2$ is a vital indicator of safety and an outcome measure of efficacy. $SpO_2$ was obtained for all subjects during the baseline week and was continuously monitored throughout all treatment visits using a finger probe (Capnostream™ 35 Portable Respiratory Monitor). Values were obtained each second to generate a times-series. Exponential smoothing at an appropriate dampening factor (1 – α) was carried out on the raw data. This was done to remove high frequency noise within the datasets and improve pattern recognition during analysis.

Figure D1 shows a typical time series graph of $SpO_2$ collected during a patient treatment visit. The blue line shows the smoothed SpO2 values for a sequence of three 2-minute doses of S-1226 (12%). The vertical broken lines indicate the 2-minute intervals used in the analyses. These include a baseline, the 2-minute treatment period and 2 minutes post treatment.

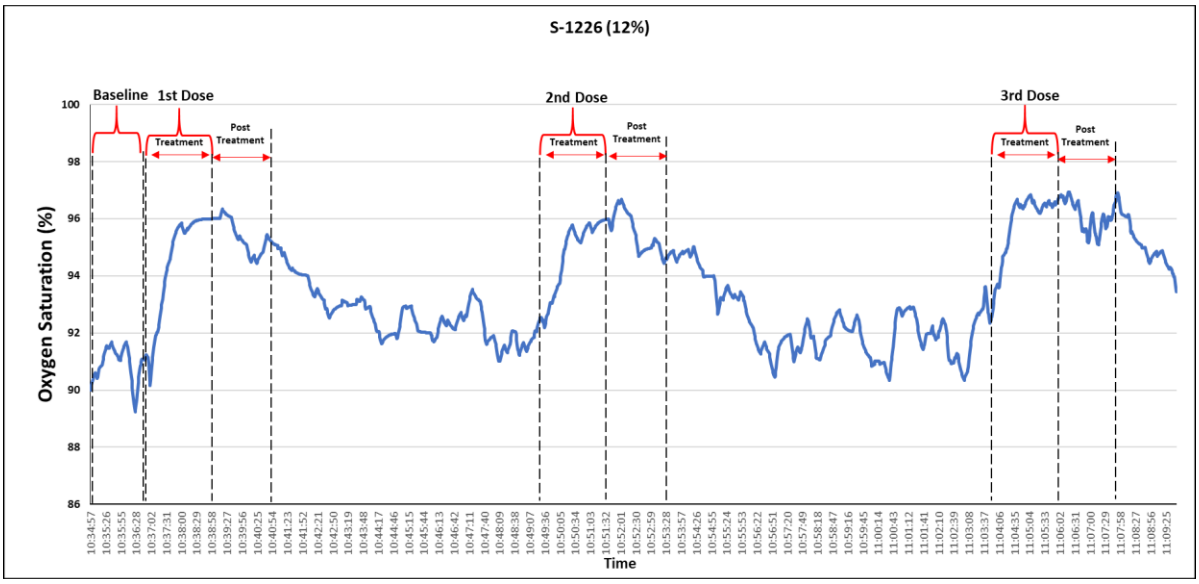


**Supplemental Figure D1: Smoothed Oxygen Saturation ($SpO_2$) graph for CFT01 given S-1226 (12%).** This figure illustrates the oxygen saturation for one CF Subject during the dose escalation week of the clinical trial. The vertical broken lines show the $SpO_2$ at baseline, during the 2-minute treatment period with S-1226 and for 2 minutes after treatment. There is a rapid increase in $SpO_2$ with each successive dose, with no evidence of tachyphylaxis (tolerance). This was a typical response for all subjects. No subject showed a decrease in their $SpO_2$ for any treatment. Data for all subjects are shown in Table 5 and Figure 4.

**Supplemental Table D1: Summary of Oxygen Saturation ($SpO_2$) following treatment all Six CF Patients.**

| | CFT01 | CFT02 | CFT03 | CFT04 | CFT05 | CFT07 | Averages | SD |
|---|---|---|---|---|---|---|---|---|
| **Baseline** | | | | | | | | |
| SpO2 (%) | 93.01 | 95.65 | 91.71 | 93.00 | 96.00 | 95.88 | 94.21 | 1.86 |
| **S1226 (4%)** | | | | | | | | |
| SpO2 (%) Before | 94.66 | 95.10 | 92.24 | 92.40 | 95.04 | 95.34 | 94.13 | 1.42 |
| SpO2 (%) During Treatment | 95.35 | 95.75 | 94.25 | 93.92 | 97.77 | 96.71 | 95.62 | 1.46 |
| SpO2 (%) Post Treatment | 94.82 | 96.07 | 96.07 | 94.19 | 96.93 | 97.51 | 95.93 | 1.25 |
| **S1226 (8%)** | | | | | | | | |
| SpO2 (%) Before | 92.50 | 95.52 | 91.42 | 92.84 | 96.50 | 95.11 | 93.98 | 2.00 |
| SpO2 (%) During Treatment | 94.93 | 96.28 | 93.87 | 93.81 | 97.70 | 96.24 | 95.47 | 1.54 |
| SpO2 (%) Post Treatment | 95.27 | 97.29 | 94.96 | 94.40 | 97.59 | 96.96 | 96.08 | 1.36 |
| **S1226 (12%)** | | | | | | | | |
| SpO2 (%) Before | 92.55 | 95.56 | 92.63 | 92.93 | 96.18 | 95.37 | 94.20 | 1.67 |
| SpO2 (%) During Treatment | 95.94 | 97.40 | 94.88 | 93.78 | 98.64 | 96.41 | 96.17 | 1.74 |
| SpO2 (%) Post Treatment | 95.86 | 97.66 | 96.10 | 94.12 | 98.77 | 96.35 | 96.47 | 1.60 |
| **Daily Dosing** | | | | | | | | |
| SpO2 (%) Before | 94.38 | 94.83 | 91.60 | 93.39 | 96.87 | 94.84 | 94.32 | 1.75 |
| SpO2 (%) During Treatment | 95.97 | 96.41 | 94.23 | 93.92 | 97.52 | 96.75 | 95.80 | 1.43 |
| SpO2 (%) Post Treatment | 95.52 | 96.73 | 95.22 | 94.36 | 97.66 | 96.57 | 96.01 | 1.19 |
| **Home Extension** | | | | | | | | |
| SpO2 (%) Before | Not Applicable | | | | | 95.12 | 95.12 | N/A |
| SpO2 (%)During Treatment | | | | | | 96.79 | 96.79 | N/A |
| SpO2 (%) Post Treatment | | | | | | 96.05 | 96.05 | N/A |
| **Follow up (Day after final Treatment)** | | | | | | | | |
| SpO2 (%) | 93.50 | 96.00 | 91.00 | 95.00 | 96.50 | 94.24 | 94.37 | 1.99 |
| **Follow up (10-14 days after final treatment)** | | | | | | | | |
| | 96.50 | 98.00 | 92.5 | N/A | 98.00 | 94.84 | 95.97 | 2.34 |

Table D1 shows that $SpO_2$ values at baseline were below the normal range, ($SpO_2$ ≤94%), for 3/6 subjects. Treatment with S-1226 resulted in a return to a normal $SpO_2$>94% for CFT01, CFT03 and CFT04 and an improvement in $SpO_2$ for CFT02, 05 and 07. There was a rapid and relatively short-term (15 min) increase in $SpO_2$ in all subjects ranging from 2-3% during and following treatment. Statistical analysis showed a significant increase in $SpO_2$ post treatment when compared to their respective before treatment values (p value <0.05) for all subjects. There was no evidence of tachyphylaxis (tolerance). In fact, there appeared to be a general upward trend in $SpO_2$ with each successive treatment, suggesting a cumulative beneficial effect.

### Supplemental E: Spirometry and Lung Clearance Index (LCI)

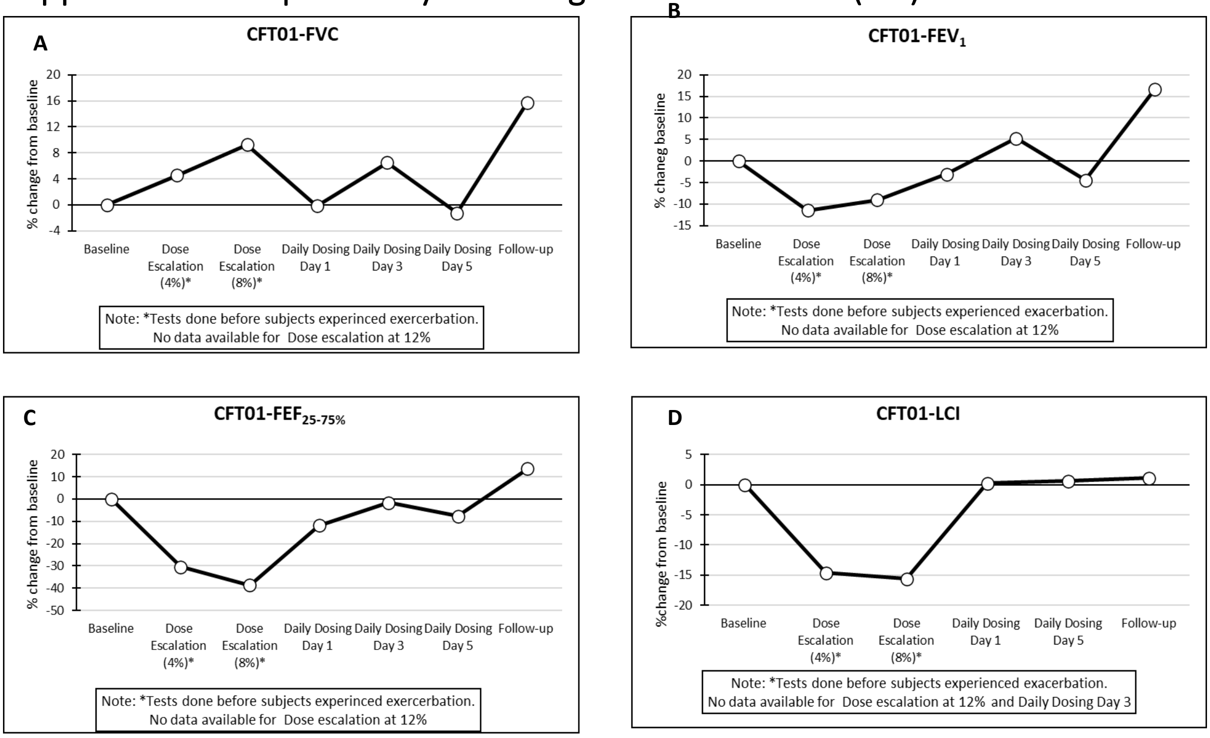


**Supplemental Figure E1: Spirometry and LCI Results for CF Subject CFT01. E1-A shows %change in Forced vital capacity (FVC); E1-B %change in Forced Expiratory volume in 1 second ($FEV_1$); E1-C %change in Forced Expiratory Flow at 25-75% ($FEF_{25-75\%}$); E1-D % improvement in Lung Clearance Index (LCI) from baseline.** Subject was dosed with S-1226 (4, 8 and 12% $CO_2$) during the dose escalation week. Subject then received S-1226 (12% $CO_2$) twice daily for 5 consecutive days during the daily dosing study. All values above the line represent improvements in function. The effect of S-1226 was greatest for FVC and $FEV_1$.

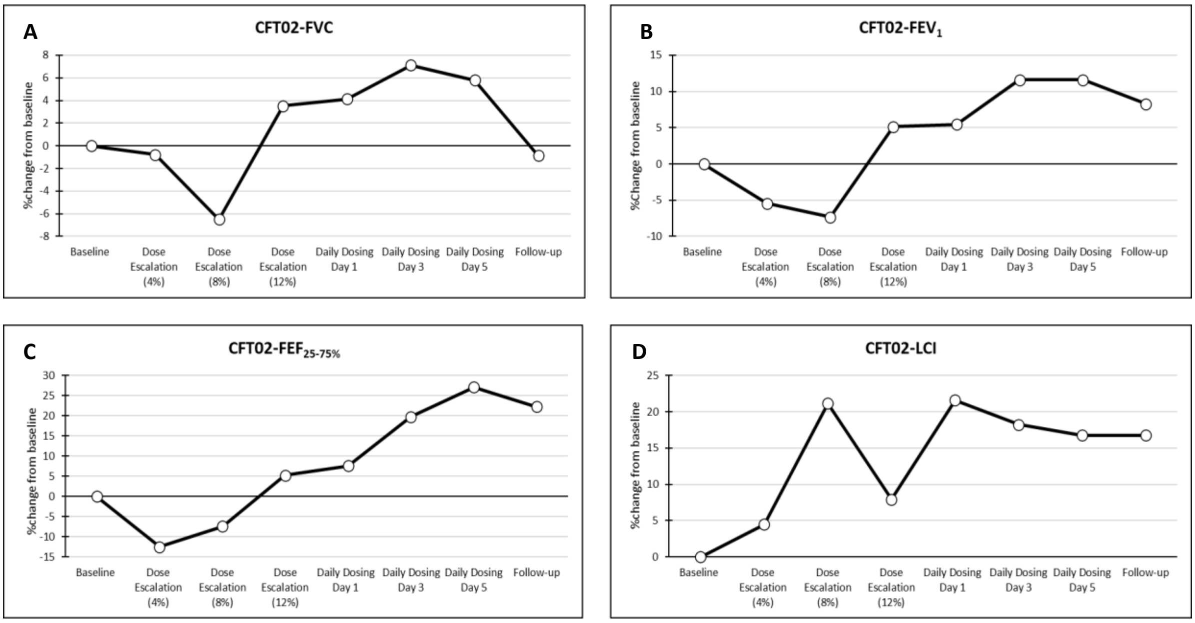


**Supplemental Figure E2: Spirometry and LCI Results for CF Subject CFT02. E2-A shows %change in Forced vital capacity (FVC); E2-B %change in Forced Expiratory volume in 1 second ($FEV_1$); E2-C %change in Forced Expiratory Flow at 25-75% ($FEF_{25-75\%}$); E2-D % improvement in Lung Clearance Index (LCI) from baseline**. Subject was dosed with S-1226 (4, 8 and 12% $CO_2$) during the dose escalation week. Subject then received S-1226 (12%

$CO_2$) twice daily for 5 consecutive days during the daily dosing study. All values above the line represent improvements in function. Improvement in LCI during the dose escalation and daily dosing proved to be greatest measure of S-1226 effect.

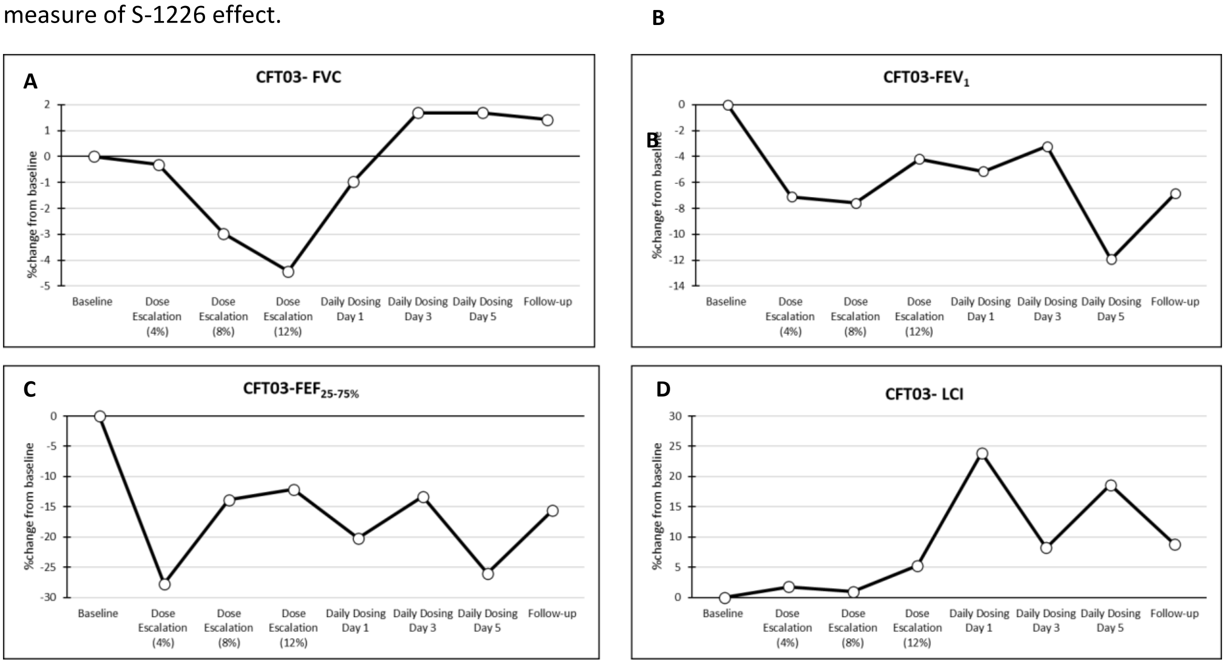


**Supplemental Figure E3: Spirometry and LCI Results for CF Subject CFT03. E3-A shows % change in Forced vital capacity (FVC); E3-B %change in Forced Expiratory volume in 1 second ($FEV_1$); E3-C % change in Forced Expiratory Flow at 25-75% ($FEF_{25-75\%}$); E3-D % improvement in Lung Clearance Index (LCI) from baseline.** Subject was dosed with S-1226 (4, 8 and 12% $CO_2$) during the dose escalation week. Subject then received S-1226 (12% $CO_2$) twice daily for 5 consecutive days during the daily dosing study. All values above the line represent improvements in function. Improvement in LCI during the dose escalation and daily dosing proved to be greatest measure of S-1226 effect.

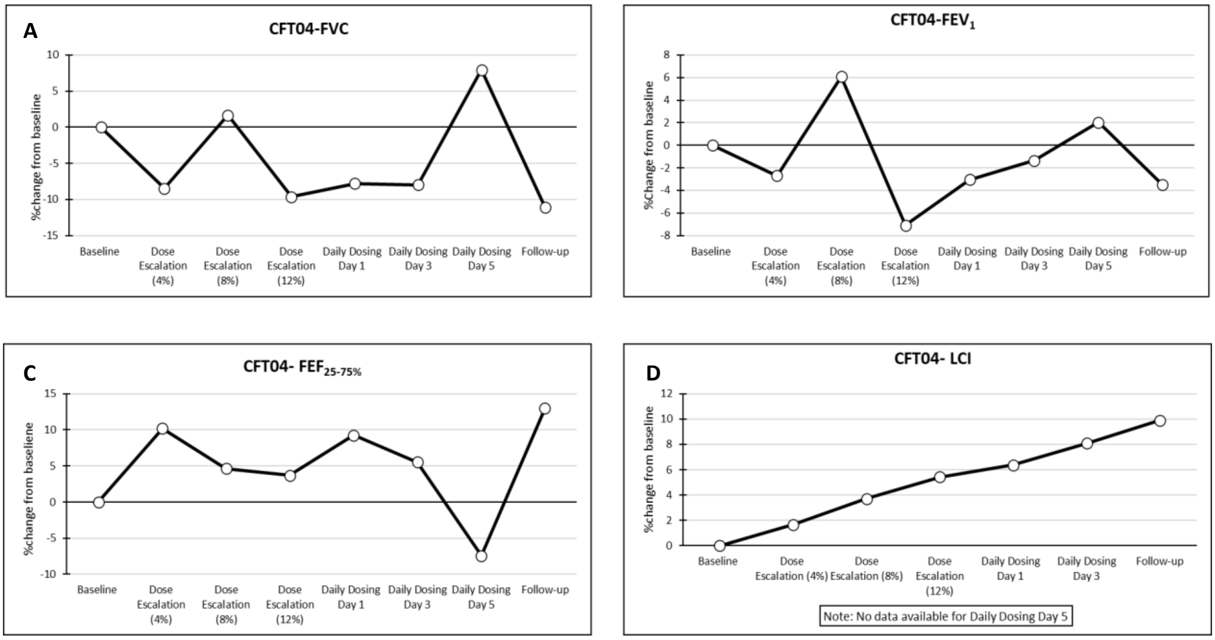


**Supplemental Figure E4: Spirometry and LCI Results for CF Subject CFT04. E4-A shows % change in Forced vital capacity (FVC); E4-B %change in Forced Expiratory volume in 1 second ($FEV_1$); E4-C %change in Forced Expiratory Flow at 25-75% ($FEF_{25-75\%}$); E4-D % improvement in Lung Clearance Index (LCI) from baseline.** Subject was dosed with S-1226 (4, 8 and 12% $CO_2$) during the dose escalation week. Subject then received S-1226 (8% $CO_2$) twice daily for 5 consecutive days during the daily dosing study. All values above the line represent improvements in function. Improvement in LCI over the course of treatment for subject CFT04 showed the greatest step wise effect of S-1226. This subject also produced copious sputum during treatments, see figures G1 and G2.

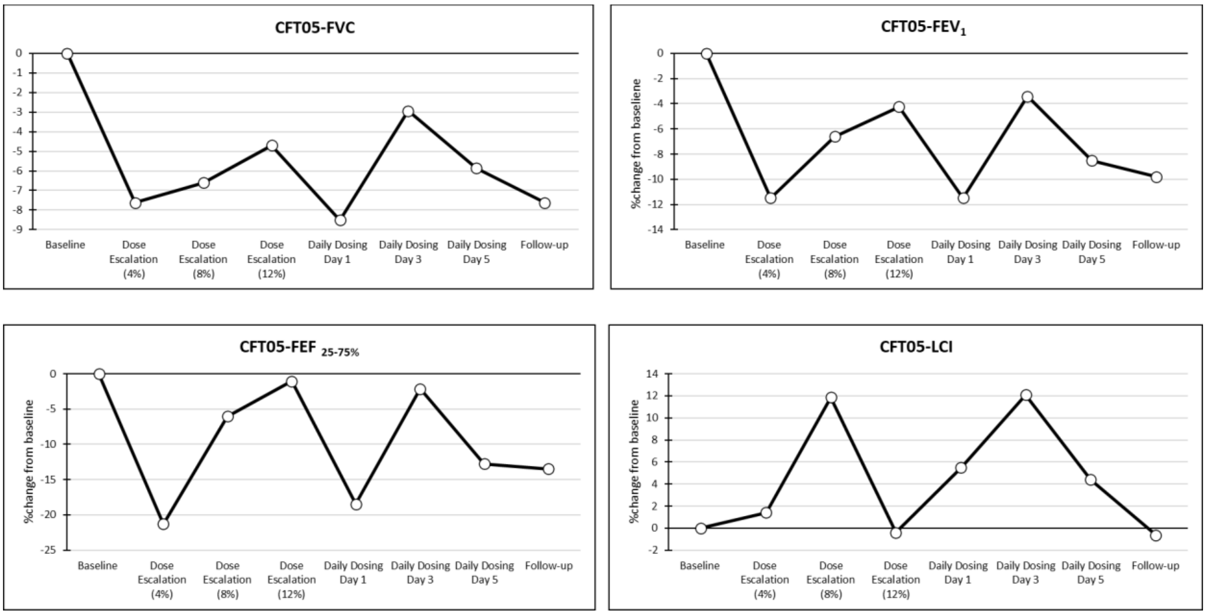


**Supplemental Figure E5: Spirometry and LCI Results for CF Subject CFT05**. **E5-A shows %change in Forced vital capacity (FVC); E5-B %change in Forced Expiratory volume in 1 second ($FEV_1$); E5-C %change in Forced Expiratory Flow at 25-75% ($FEF_{25-75\%}$); E5-D % improvement in Lung Clearance Index (LCI) from baseline.** Subject was dosed with S-1226 (4, 8 and 12% $CO_2$) during the dose escalation week. Subject then received S-1226 (12% $CO_2$) twice daily for 5 consecutive days during the daily dosing study. All values above the line represent improvements in function**.** The LCI improved from baseline in dose ranging and daily dosing. Two values during treatments reached normal values: 7.34 and 7.32 (Normal range 5.6-7.5). Th LCI returned to baseline values at follow-up.

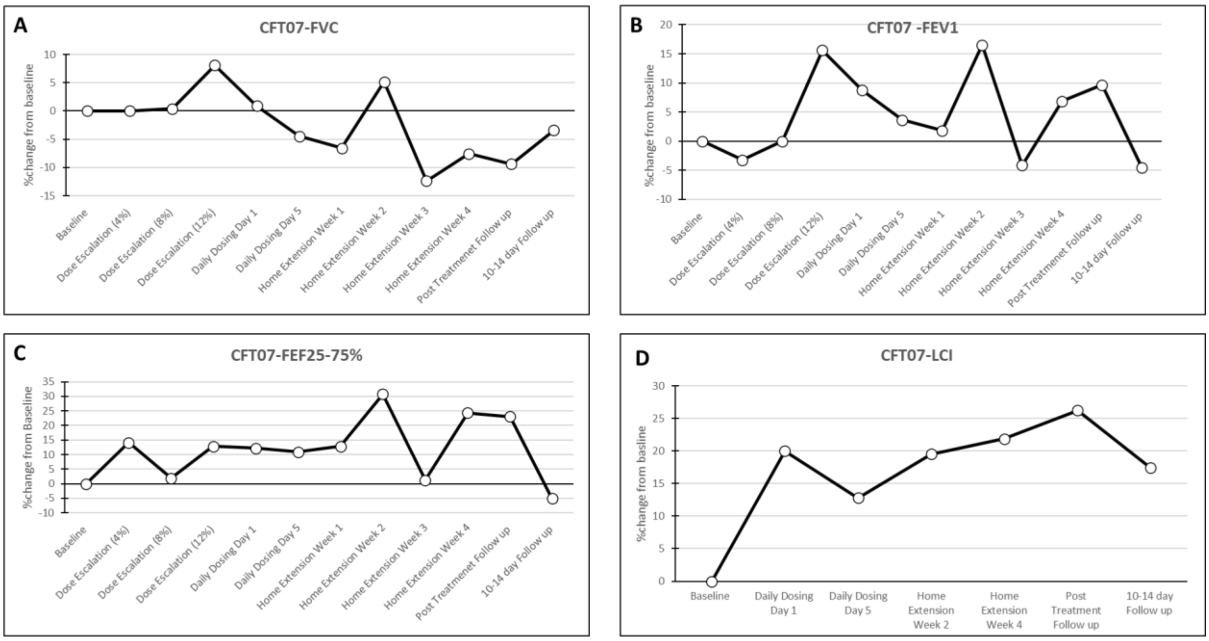


**Supplemental Figure E6: Spirometry and LCI Results for CF Subject CFT07**. **E6-A shows %change in Forced vital capacity (FVC); E6-B %change in Forced Expiratory volume in 1 second ($FEV_1$); E6-C %change in Forced Expiratory Flow at 25-75% ($FEF_{25-75\%}$); E6-D % improvement in Lung Clearance Index (LCI) from baseline.** Subject was dosed with S-1226 (4, 8 and 12% $CO_2$) during the dose escalation week. Subject then received S-

1226 (12% $CO_2$) twice daily for 5 consecutive days during the daily dosing study. Subjects continued to receive two doses of S1226 twice daily during the at home study extension for 4 weeks. No doses were missed. LCI values (E6-D) consistently improved form baseline and was sustained 10-14 days following treatment. Change in FVC, $FEV_1$, $FEF_{25-75\%}$ values varied considerably with no clear pattern emerging.

#### Supplemental F: Sputum collection

**Figure F1: Change in daily sputum weights** compared to baseline during treatment with S-1226 (4-12%) for

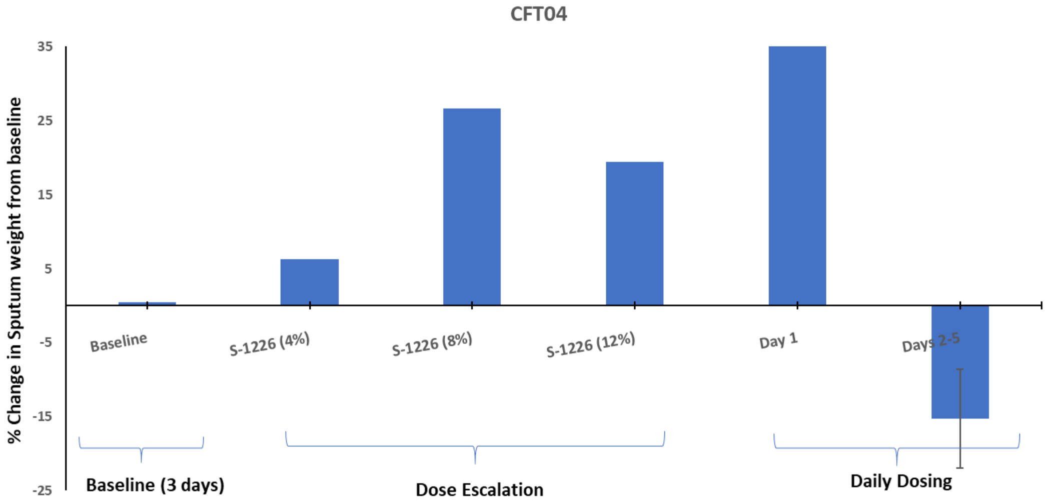


subject CFT04.The expectorated sputum from subject CFT04 was yellow coloured and thick (Figure F2).

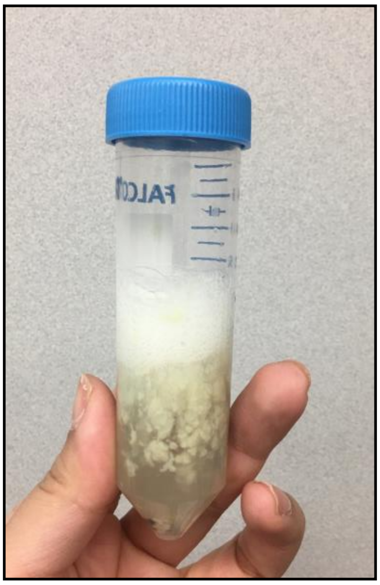

**Mucus expectorated by subject CFT04, 20 minutes after S-1226 (8%) treatment.** Note the large clumps. This may suggest that the mucous is coming from deep in the lung, probably from an area with bronchiectasis.

**Supplemental Figure F2: Mucus expectorated by subject CFT04 immediately after S-1226 (8%) treatment.** The expectorated mucous following S-1226 treatment was more liquid and less adhesive to the walls of the collection tubes compared to baseline sputum samples Video available on request.

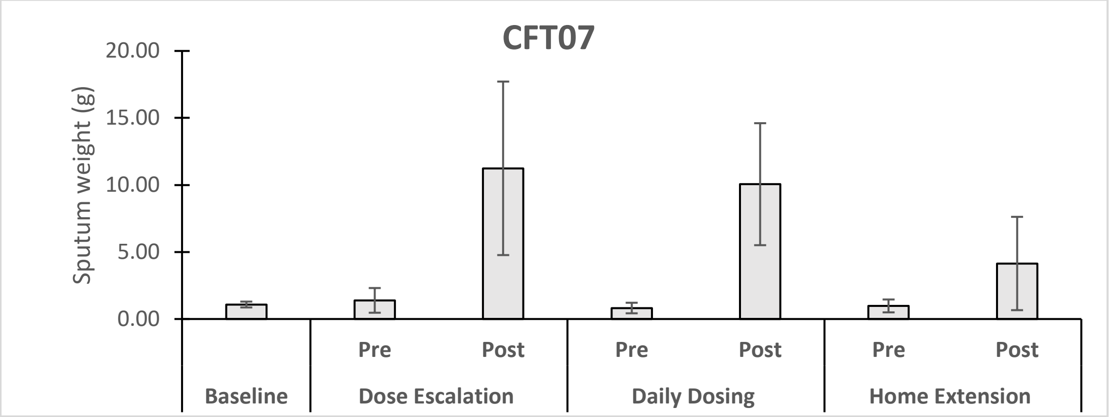


**Supplemental Figure F3: Mucus expectorated by CFT07 before (pre) and 2 hours after (post) treatment with S-1226**. Sputum weight (g) peaked during the first week of treatment but began to decline back to baseline by the home extension treatment weeks. Subject consistently produced more quantities of sputum 2 hour following treatment, a pattern also observed in CFT02 and CFT04.
